# Prognostic significance of protein-coding and long non-coding RNA expression profile in T-cell acute lymphoblastic leukemia

**DOI:** 10.1101/2021.05.06.21255823

**Authors:** Deepak Verma, Shruti Kapoor, Disha Sharma, Jay Singh, Gunjan Sharma, Sarita Kumari, Mohit Arora, Sameer Bakhshi, Rachna Seth, Baibaswata Nayak, Atul Sharma, Raja Pramanik, Jayanth Kumar Palanichamy, Sridhar Sivasubbu, Vinod Scaria, Rajive Kumar, Anita Chopra

## Abstract

T-acute lymphoblastic leukemia (T-ALL) is an aggressive hematological malignancy associated with poor outcome. To unravel gene-expression profile of immunophenotypic subtypes of T-ALL, we did transcriptome analysis in 35 cases. We also analyzed the prognostic relevance of 23 targets: protein-coding genes, histone modifiers and long non-coding RNA (lncRNA) expression profile, identified on RNA sequencing, on an independent cohort of 99 T-ALL cases. We found high expression of *MEF2C* to be associated with prednisolone resistance (*p=*0.048) and CD34 expression (*p*=0.012). *BAALC* expression was associated with expression of CD34 (*p=*0.032) and myeloid markers (*p=*0.021). *XIST* and *KDM6a* expression levels were higher in females (*p=*0.047 and 0.011, respectively). Post-induction minimal residual disease (MRD) positivity was associated with high lncRNA PCAT18 (*p=*0.04), *HHEX* (*p=*0.027) and *MEF2C* (*p=*0.007). Early thymic precursor (ETP-ALL) immunophenotype was associated with high expression of *MEF2C* (*p=*0.003), *BAALC* (*p=*0.003), *LYL1* (*p=*0.01), *LYN* (*p=*0.01), *XIST* (*p=*0.02) and low levels of *ST20* (*p=*0.007) and *EML4* (p=0.03). On survival analysis, *MEF2C* high expression emerged as significant predictor of 3-year event free survival (EFS) (low 71.78±6.58% vs high 36.57±10.74%, HR 3.5, *p=*0.0003) and overall survival (OS) (low 94.77±2.96% vs high 78.75±8.45%, HR 4.88, *p=*0.016) in our patients. LncRNA MALAT1 low expression also emerged as predictor inferior OS (low 76.02±10.48 vs high 94.11±3.31, HR 0.22, *p=*0.027).

## Introduction

T-lineage acute lymphoblastic leukemia (T-ALL) accounts for ∼25% of adults and 10%–15% of pediatric ALL cases^1^. In the past several years immunological, cytogenetic and genomic approaches have been extensively utilized for a better understanding of the genomic organization of functional elements and their alterations in leukemia. This, combined with techniques of cell biology, applied to T-ALL has led to major advances in our understanding of this disease and has allowed the development of novel therapeutic approaches.

The malignant transformation that culminates in T-ALL is a multi-step process, in which both genetic and epigenetic alterations of key cellular pathways coordinate to produce the T-ALL phenotype (Peirs et al., 2015; Girardi et al., 2017). T-ALL pathogenesis is characterized by an abnormality with highly variable gene expression profiles. The genetic alterations and transcriptomic signatures are often used for classification of T-ALL patients into molecular subtypes exhibiting distinct clinical outcomes which are partly determined by their underlying oncogenic signalling pathways, such as *IL7R/JAK/STAT, PI3K/AKT*^5-7^ and *RAS/MEK/ERK*^8^ signaling. Aberrant expression of a diverse group of transcription factors such as *LYL1, LMO1, LMO2, TAL1, TLX1, TLX3, HOXA, NKX2*.*1, NKX2*.*2, NKX2*.*5, MYC, MYB* and *SPI1* has been reported in distinct T-ALL subtypes^3,4^. Some of the less understood facets in T-ALL include epigenetic deregulation, ribosomal dysfunction, and altered expression of oncogenic miRNAs and long non-coding RNA. Apart from the genetic alterations, epigenetic alterations such as DNA methylation, histone modifications, chromosomal remodelling are chromosomal topology have also been shown to be involved in T-ALL pathogenesis and modulation of clinical response (Van der Meulen et al., 2014; Kloetgen et al., 2020).

More recently, a greater understanding of molecular pathophysiology and immunophenotyping methods led to the refinement of classification of T-ALL but the clinical relevance of these subtypes remains controversial^1,9-11^. The only subtype of T-ALL which has got a place in the 2016 revision of WHO, is early thymic precursor ALL (ETP-ALL)^11,12^. In this study, we analyzed the relevance of T-ALL immunophenotyping and the expression pattern of protein-coding genes, epigenetic modifiers and long non-coding RNAs in T-ALL classification and also determined their association with patient prognosis.

## Materials and Methods

A total of 134 T-ALL cases, newly diagnosed based on morphology, cytochemistry and immunophenotyping, were enrolled in this study. These cases were immunophenotypically classified into immature (pro T- and pre T-), cortical and mature T-ALL based on the EGIL criteria^13^. ETP-ALL was recognized based on the previously defined criteria^12^. The Patients were divided into 2 cohorts: discovery (n=35) and validation cohort (n=99). Total RNA sequencing was done in the discovery cohort. The clinical and prognostic relevance of transcriptomic features identified in the discovery cohort were tested in the validation cohort. All patients or guardians gave informed consent for blood/bone marrow collection and biological analyses, in agreement with the Declaration of Helsinki. The study was approved by the Institutional Ethical Committee, All India Institute of Medical Sciences, New Delhi, India. Transcriptome data of pooled RNA extracted from 5 normal human thymus samples were used as a non-cancer control (kind courtesy Dr Jan Cools, Belgium).

### Discovery cohort

In the discovery cohort (n=35), there were 13 immature (including 5 ETP-ALL), 17 cortical and 5 mature T-ALL cases with a median age of 14 years (range,1 to55 year). There were 26 males (19 pediatrics, 7adults) and 9 females (7 pediatrics, 2 adults).

### RNA sequencing and analysis of the discovery cohort

RNA was extracted from freshly isolated patient blood samples by the TRIzol method (Thermo Fisher Scientific, Massachusetts, USA). Transcriptome data of pooled total RNAs from 5 normal human thymus was used as a control (kindly gifted by Dr Jan Cools, Belgium) for analysis of epigenetic modifiers and miRNA. Paired-end whole transcriptome sequencing was performed on the Illumina HiSeq2000 platform using Truseq RNA sample preparation kit (Illumina, San Diego, California, U.S.). Sequence reads were processed to identify the expression profile of protein-coding and non-coding RNAs by supervised and unsupervised approaches **(supplementary methods)**. To investigate the role of histone modifiers in T-ALL development, transcript abundance of epigenetic modifiers was measured across mature, cortical, and immature subtypes of T-ALL (supplementary methods). To know the novel lncRNA transcript, a computational pipeline combining Open Reading Frame (ORF) prediction coupled with coding potential calculator (CPC algorithm) to annotate the *protein-coding* potential of transcripts, was used (supplementary methods). Biomart was utilized to identify the annotated miRNA and transcripts.

Transcripts with ≥ 2-FC were selected as differentially expressed putative coding and non-coding transcripts in the given subgroup and heatmap were plotted using MeV. Gene Ontology enrichment and KEGG pathway analyses were used to explore the potential biological processes, cellular components, and molecular functions of differentially expressed genes (DEG). Gene functional analysis was done on DAVID 6.8 database^14,15^. To predict the functions of highly overexpressed validated genes for the protein-coding genes, histone modifiers, lncRNA in all three subgroups of T-ALL, a correlation network analysis was performed using the Rcorr package in R based on Spearman correlation and visualized using Cytoscape^16^. Detailed methods are described in the supplementary methods.

### Validation cohort

Candidate genes found differentially expressed in the discovery cohort were further validated through real-time PCR in an independent cohort of 99 {46 immature (including 15 ETP-ALL), 43 cortical, 10 mature} T-ALL cases including 70 pediatrics (61 males, 9 females), 29 adults (25 males, 4 females). The age of the patients ranged from 3 to 65 years with a median age of 12 years. The expression levels of 23 targets: *BAALC, HHEX, MEF2C, FAT1, LYL1, LMO2, LYN, TAL1, DOT1L, XIST, PCAT18, PCAT14, LNC202, LNC461, LNC648, MEF2C-AS1, ST20, RAG, EP300, EML4, EZH2, MALAT1* and *KDM6A* were compared with patient characteristics and survival. (Supplementary Methods). The relative quantitation method by real-time PCR was used to calculate the fold change in gene expression relative to housekeeping gene ABL1. Consistently selected genes from discovery and validation cohort followed the similar distribution of expression. Detailed experimental plan used to study the discovery and validation cohorts is mentioned in Supplementary Methods.

### Treatment

In the discovery cohort, 14 patients were treated with ICICLE protocol, 3 with Berlin-Frankfurt-Munster-90 (BFM-90) protocol, 1 with INCTR, 1 with Holzer’s protocol and 3 with hyper CVAD. 13 patients did not receive treatment/succumbed to the disease before taking treatment.

In the validation cohort, seventy-one patients were treated with ICICLE protocol, 15 with Berlin-Frankfurt-Munster-90 (BFM-90) protocol and 3 with Hyper CVAD. Ten patients did not take any treatment. Two patients died during induction chemotherapy. Complete remission was defined as bone marrow blasts <5% with a recovery of blood counts at the end of 4 weeks of induction chemotherapy. Any failure to do so (including the persistence of leukemic blasts in an extramedullary site), or death during induction therapy due to any cause, was considered as induction failure. Patients who failed with one protocol were re-induced with another.

## Statistical analyses

Fisher’s exact test for categorical data and the nonparametric Mann-Whitney U for continuous variables were used to compare baseline clinical variables across groups in the validation cohort. A P-value ≤0.05 (two-sided) was considered significant. Event-free survival (EFS) was defined as the time from diagnosis to the date of the last follow-up in complete remission or the first event (i.e., induction failure, relapse, secondary neoplasm, or death from any cause). Failure to achieve remission due to non-response was considered an event at time zero. Survival was defined as the time from diagnosis to death or the last follow-up. Patients lost to follow-up were censored at the last contact. The last follow up was carried out on April 2020. The Kaplan-Meier method was used to estimate survival rates, with the differences compared using a two-sided log-rank test. Univariate and multivariate Cox proportional hazard models were constructed for EFS and OS. Covariates included sex, WBC (<50×10^9^/L, ≥50×10^9^/L), age (<12 years vs. ≥12 years), gene expression, immunophenotype and response to prednisolone treatment and presence of minimal residual disease after the end of induction chemotherapy. Patients with high and low expression were delineated using maximally selected rank statistics as implemented in the maxstat R package (http://cran.r-project.org/web/packages/maxstat/index.html) for each target (*BAALC, HHEX, MEF2C, FAT1, LYL1, LMO2, LYN, TAL1, DOT1L, XIST, PCAT18, PCAT14*, LINC00202, LINC00461, LINC00648, *MEF2C-AS1, ST20, RAG1, EP300, EML4, EZH2, MALAT1* and *KDM6A)*. Statistical analyzed were performed using the SPSS statistical software package (version 20.0), STATA software (version 11) and R statistical software (version).

## Data Availability

The RNA-Seq raw data that support the findings of this study are available from the corresponding author upon reasonable request.

## RESULTS

### A. Discovery cohort

#### Distinct profiling of differentially expressed genes expression among T-ALL immunophenotype

Comparison of gene expression profile among the three different subtypes defined by immunophenotyping revealed a total of 2,318 genes to be differentially expressed (Figure 2a) (supplementary1-4). In **immature T-ALL** subtype, transcription factors which control early hematopoiesis, such as *MEF2C, TP63, HHEX, RUNX2, HOXA10, HOXA9, RUNX1T1* and *ZBTB16*; homeobox gene like *HOPX*; *LYL1, LMO2* and *LYN (a* tyrosine kinase coding gene) were highly expressed. Interestingly, LMO2 was previously reported to be expressed under the presence of LYL1 gene only but in our data LMO2 was found upregulated in the absence of significant overexpression of *LYL1, a* gene reported to be critical for oncogenic functions of LMO2. *BAALC* and *MN1* genes, previously reported in AML^17^, were overexpressed. *BAALC* associated genes like *IGFBP7* and *PROM1* (CD133), which are known to confer chemoresistance, were also overexpressed^18^. Also, *MAML3, NT5E* (CD73) and *ARID5B* had significant overexpression in immature T-ALL. Moreover, genes not previously described in T-ALL like PLD4 and TP63, were overexpressed. In **cortical T-ALL**, the cortical thymocytes - defining *CD1A* gene was overexpressed. Homeobox domain genes, *NKX2-1, TLX1, TLX3*, involved in T-cell development were overexpressed. Also, *FAT1, FAT3, RAG1, EREG, CD1C, AKAP-2, IL-4, PRTG, TCL-6, ZP1* and *TRAV* genes were overexpressed. Comparison of CD1a+/sCD3+ and CD1a+/sCD3-groups, revealed that *TLX1*, in contrast to reported literature (Ferrando 2002), was overexpressed in the former group whereas *TLX3* was overexpressed in the latter. In **mature T-ALL**, *APC2, BCL3, CCR4, CDKN2A, EML4, HIST1H4G, HIST2H2B, NCOR2, ST20* and *TRAV22* genes were overexpressed (figure 2a). *TAL1* expression was seen in T-ALL cases with mature immunophenotype. A complete list of DEG is shown in the supplementary file.

**Figure 1:**
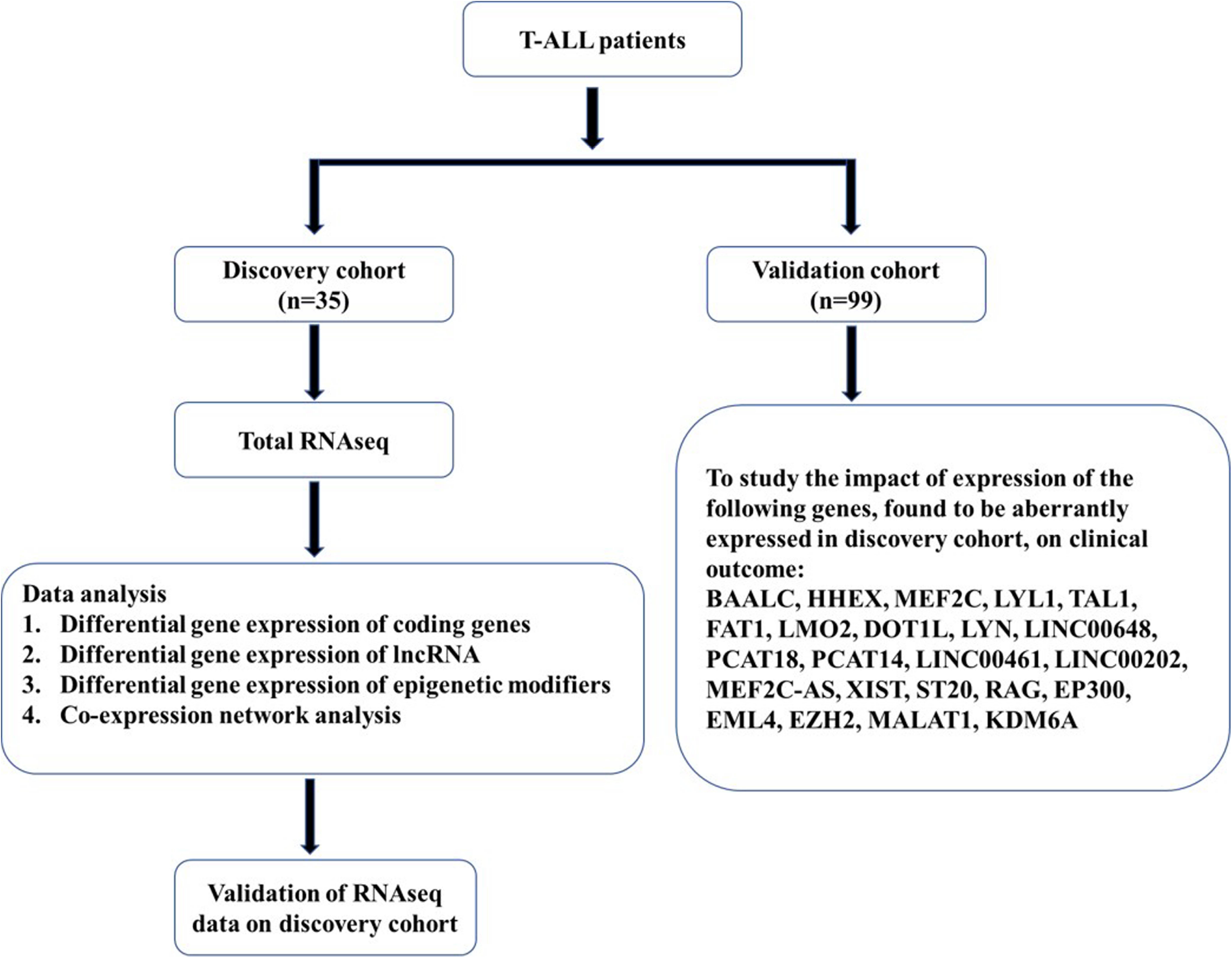
Workflow for identification and validation of molecular markers in T-ALL patients.

**Figure 2:**
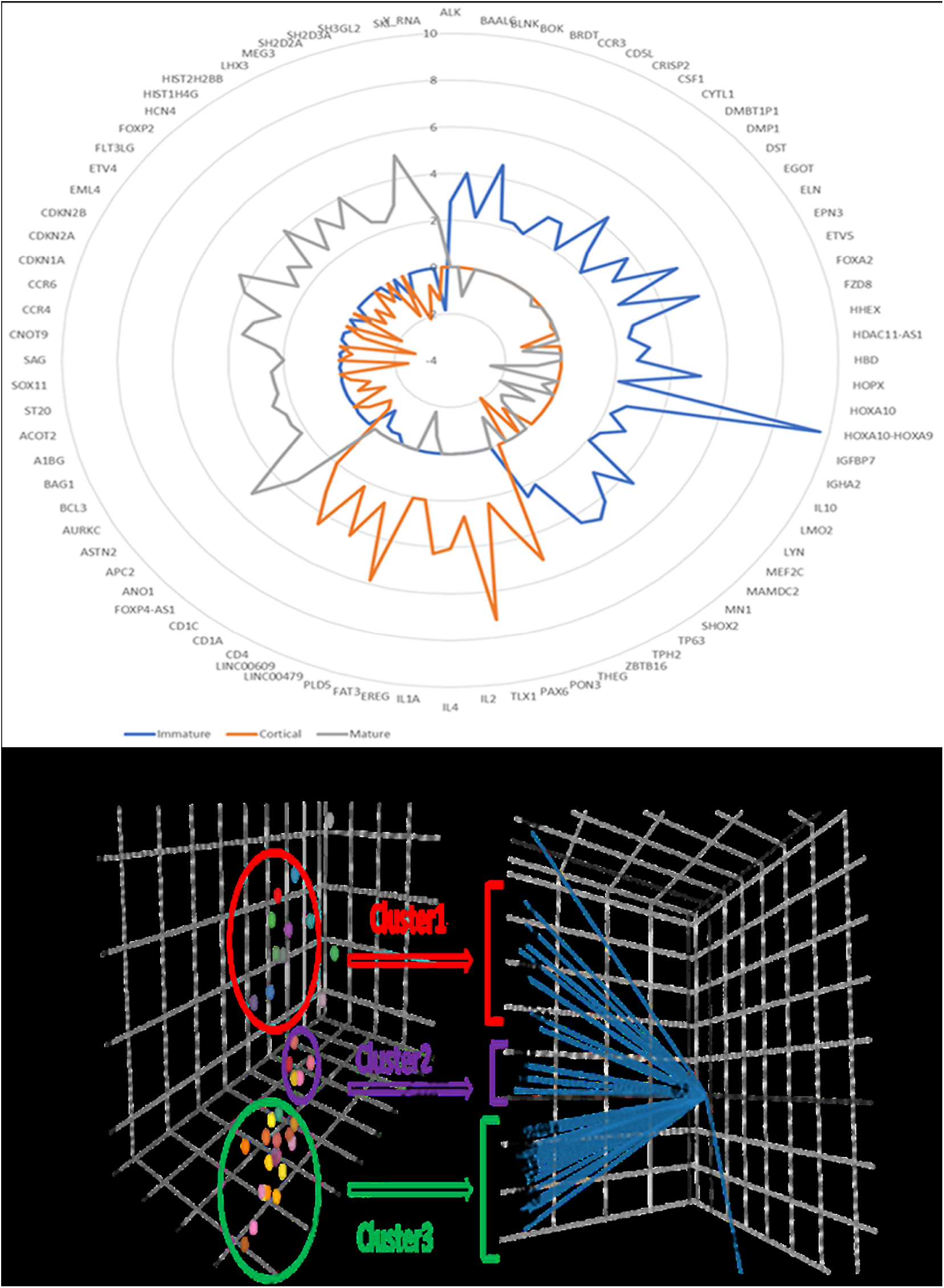
**(a)** Radar graph of Differentially expressed genes among T-ALL subgroups: immature (Blue), cortical (Green), mature (Gray). each circle represents the fold change for the differentially expressed genes. **(b)** PCA analysis of Gene expression: Figure depicts the clustering of samples in 3 major clusters according to their normalized FPKM count. Cluster 1 comprises of 9, Cluster 2 comprises 5 and Cluster 3 comprises 19 T-ALL samples.

#### Unsupervised immunophenotype analysis segregates mixed T-ALL gene expression profile into three distinct clusters

Principal component analysis (PCA) was performed to determine variabilities in gene expression profiles (GEPs) in T-ALL concerning normal thymus tissue (kind courtesy, Dr Jan Cools, Belgium). We observed that these GEPs could be broadly separated in 3 major clusters (figure 2b), in which **Cluster 1, Cluster 2** and **Cluster 3** comprised 5, 9 and 19 cases, respectively. Three samples including one normal thymus were not present in any cluster. In major cluster 1, all 9 samples had immature T-ALL immunophenotype and further encompassed 2 sub-clusters consisting of 4 samples each while one remaining sample did not fall in any of the two sub-clusters. In 1^st^ sub-cluster, 3 samples had ETP-ALL *CD5*^*-*^ immunophenotype and one sample were *CD5*^*+*^ near-ETP-ALL (pre-T-ALL). In 2^nd^ sub-cluster, three samples were near-ETP-ALL (pre/pro-T-ALL) and one sample was ETP-ALL. Major cluster 2 consisted of 5 samples, all were cortical T-ALL. In major cluster 3, out of 19 samples, 3 were immature, 4 mature and the remaining were cortical T-ALL. Three samples that did not fall into any of the major clusters consisted of normal thymus, immature and mature T-ALL, respectively. Interestingly, principal component analysis placed the mixed T-ALL cases into three distinct categories of T-ALL subclasses.

#### mRNA expression profiling of epigenetic modifiers in T-ALL

Analysis of genes which are primarily involved in the epigenetic regulation revealed overexpression of *SETD2*, ATM, ASHIL, *KDM6A*, PHF6, SUZ12 and HDAC4 in all subtypes of T-ALL concerning normal thymic tissue. Furthermore, In immature T-ALL, HDAC9 and SMYD3 were overexpressed while *EZH2* was under-expressed. HDAC10 was under-expressed in cortical T-ALL. In mature T-ALL, *EP300*, PKN1, *EML4, DOT1L* were overexpressed while HDAC7 was under-expressed in immature and cortical T-ALL. (figure 3b)

**Figure 3:**
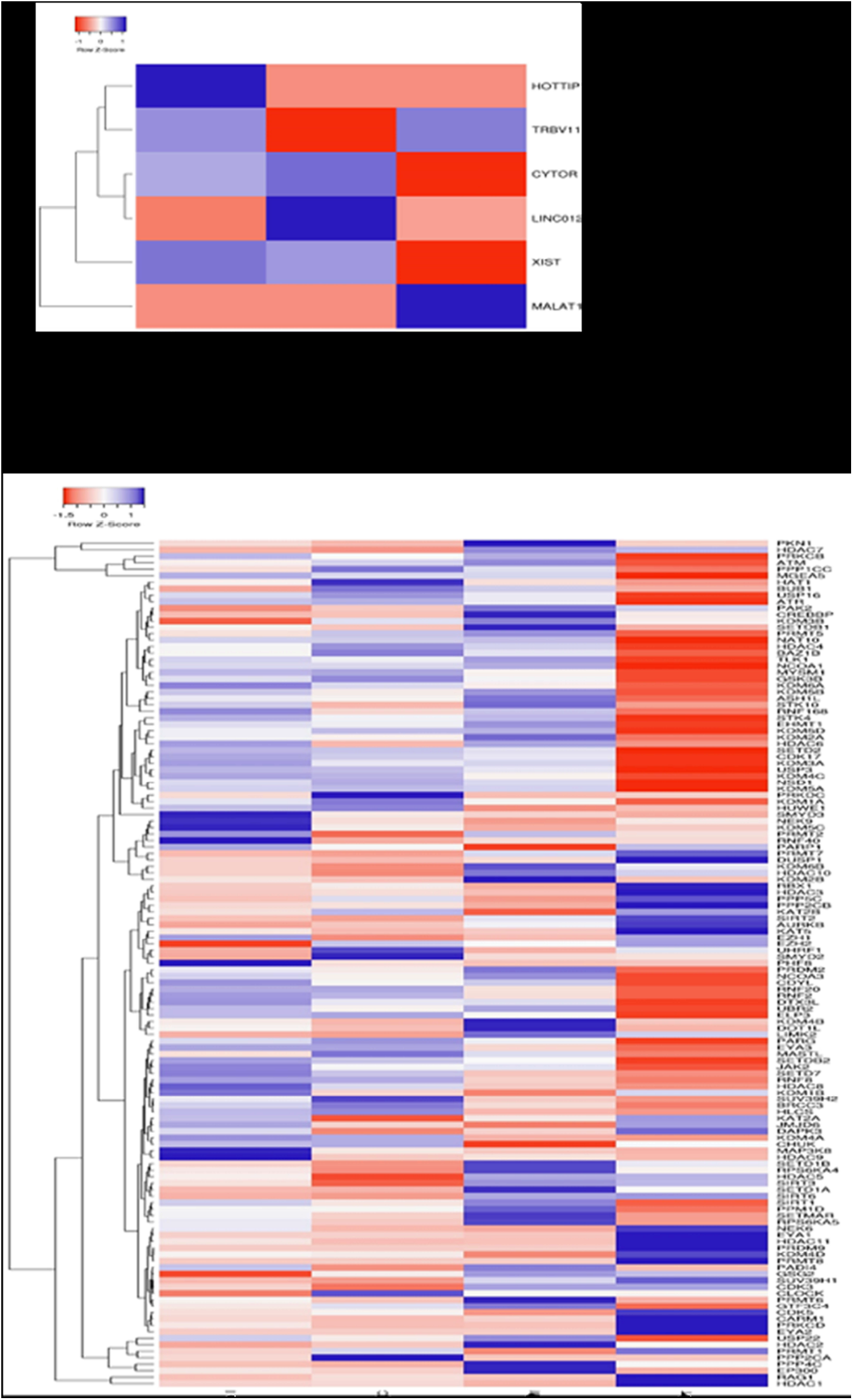
**(a)** Hierarchical clustering of differential expressed lncRNA among three T-ALL subgroups. **(b)** Hierarchical clustering differentially expressed epigenetic modifiers in T-ALL.; Each raw represents gene symbol. The “heat map” indicates high (red) or low (green) level of expression according to the scale shown.

#### Functional gene annotation and pathway analysis

To determine the biological role of DEG, which were observed during comparison of GEP in different subtypes, we performed gene ontology analysis. These DEG were strongly involved in the various biological processes including pathways involved in the inflammatory response, immune response, T cell co-stimulation, positive regulation of interferon-gamma production, signal transduction, cell-cell signaling, cell adhesion and migration. Their involvement may lead to abnormal function of various cellular components comprising plasma membrane, cell surface, extracellular space, an integral component of the plasma membrane, extracellular region, transcription factor complex, an integral component of the membrane and secretory granules. A complete list of biological processes and cell components are mentioned in figure 4a,b. We further tried to disintegrate the molecular pathways by KEGG in which the immunophenotype associated DEG could be involved. This analysis demonstrated enrichment of cellular pathways such as hematopoietic cell lineage, allograft rejection, transcriptional dysregulation in cancer, toll-like receptor signaling pathway, graft-versus-host disease, cytokine-cytokine receptor interaction, cell adhesion molecules (CAMs). Detailed pathways information showed in figure 4c.

**Figure 4:**
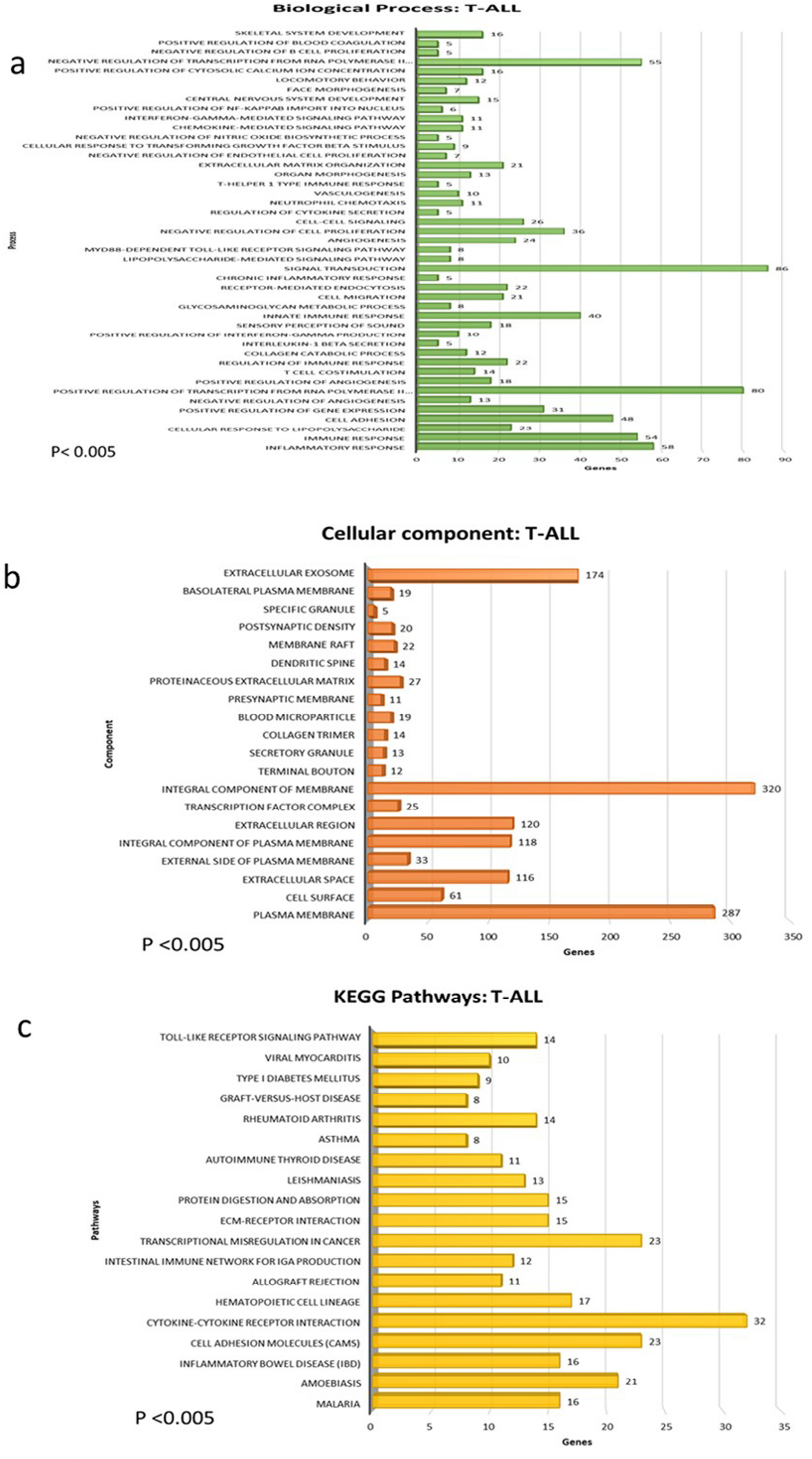
Enriched gene ontology functions of deferentially expressed genes, DAVID results the number of involved genes from differentially expressed genes in T-ALL for biological process **(a)**, cellular components **(b)** and KEGG pathways**(c)**. (p<0.005). Complete list of genes involved in the biological process and cellular component of are shown in Supplementary

#### Co-expression network analysis

Co-expression networks were constructed for differentially expressed genes between the immature, cortical and mature subgroups (with correlation score≥0.9). The genes included in the analysis were highly expressed in our RNA seq data of T-ALL patients. **Table 1** shows the relation among the *protein-coding* and *non-coding genes*. Coexpression networks of *BAALC, MEF2C*, lncRNA and epigenetic modifiers are described in supplimentry figures 1a-b, 2a-b respectively.

**Table 1:**
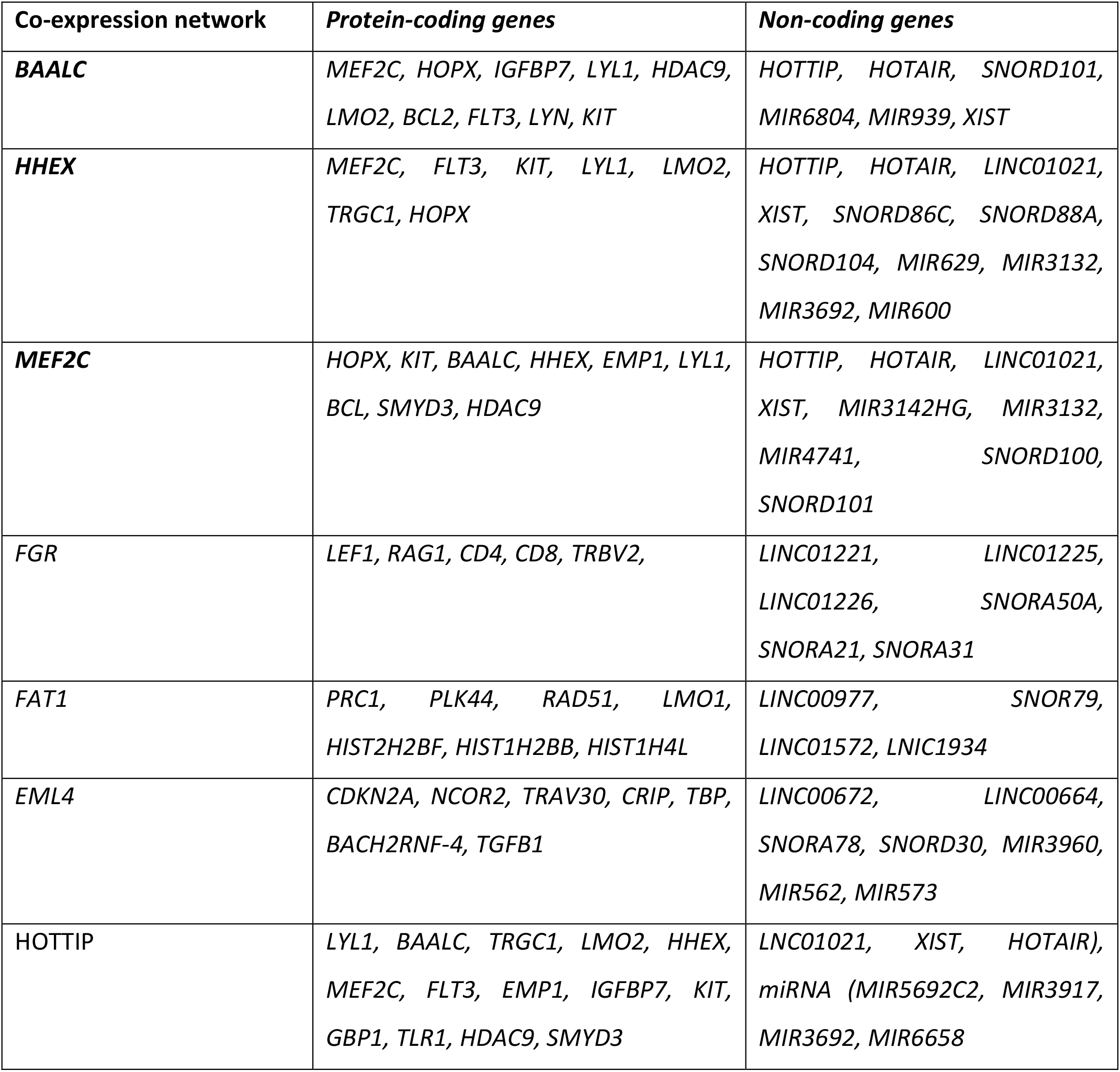
Co-expression network analysis of few selected protein-coding and non-coding genes.

### Long non-coding RNA (lncRNA)

#### RNA-seq analysis post annotation and differential expression analysis resulted in 2,243 lncRNAs which were expressed in the T-ALL samples

**A total of** 223 lncRNAs were filtered based on the criteria of >2FPKM scores. We observed differentially enriched lncRNA in the subgroup of T-ALL such as *HOTTIP* in immature T-ALL; LINC01221, LINC00202, LINC00461, LINC00648 in cortical T-ALL and *MALAT1, ST20 and TRBV11* in mature T-ALL, to be overexpressed. Interestingly, these lncRNAs have not been earlier reported in T-ALL (figure 3a). X-inactive specific transcript (*XIST)*, which is known to have a role in multiple cancers, was expressed in both immature and cortical T-ALL^19^. LUNAR1, which is known to be a specific *NOTCH1*-regulated lncRNA, was expressed in cortical and mature T-ALL^20^. The co-expression network for HOTTIP, which was highly expressed in immature T-ALL, was constructed and observed to exhibit a strong positive correlation with important transcription factors, having a role in/ previously reported to be involved in T-ALL pathogenesis. (supplimentry figure 2a). In comparison with normal thymus transcriptome, we found lncRNAs, PCAT14 and PCAT18, to be significantly overexpressed in T-ALL cases.

In addition to the annotated lncRNA though less explored in T-ALL, we also identified 1,290 novel putative *non-coding* transcripts or lncRNA in our data. All transcripts which overlapped coding potential were removed from the analysis as these could potentially contribute to false-positive annotations.

### Validation cohort

#### Correlation between gene expression, response to chemotherapy and outcome

The mRNA expression levels of differentially expressed 23 targets selected from discovery cohort based on their variable expression pattern in distinct immunophenotype, including protein-coding genes, epigenetic modifiers and lncRNA transcripts: *BAALC, HHEX, MEF2C, FAT1, LYL1, LMO2, LYN, TAL1, DOT1L, XIST, PCAT18, PCAT14, LNC202, LNC461, LNC648, MEF2C-AS1, ST20, RAG, EP300, EML4, EZH2, MALAT1* and *KDM6A*. These genes were further validated in the validation cohort to assess their clinical significance. Interestingly inconsistency to the results obtained in RNA seq analysis we observed the similar pattern of overexpression in distinct immunophenotypes in the validation cohort. High expression of *BAALC* (*p=*0.001), *MEF2C* (*p=*0.002), *LYL1* (0.018), *HHEX* (*p=*0.007) and low expression of *EZH2* (*p=*0.005) were found significantly associated with the immature T-ALL immunophenotype.

*RAG1* and FAT1 expression were higher in cortical T-ALL (*p=*0.004 and 0.033, respectively). *DOT1L* expression was higher in mature T-ALL (*p=*0.025). ETP-ALL immunophenotype was associated with high levels of *BAALC* (*p=*0.003), *MEF2C* (*p=*0.003), *LYL1* (*p=*0.01), *LYN* (*p=*0.01), *XIST* (*p=*0.02) and lower levels of *ST20* (*p=*0.007) and *EML4* (*p=*0.03). We also observed an association between CD34 positivity on immunophenotyping with expression levels of *BAALC* (*p=*0.032) and *MEF2C* (*p=*0.012). Myeloid markers (CD13/CD33) expression on immunophenotyping was associated with high *BAALC* (*p=*0.021) and low *ST20* (*p=*0.007) and low *KDM6A* (*p=*0.026). We did not find any significant association between T-ALL subtype and expression levels of PCAT14, PCAT18, *TAL1, LMO2, XIST, ST20, EP300, EML4, KDM6A*, LINC00202, LINC00461 and LINC00648. Of the 99 T-ALL patients in the validation cohort, RNA sample was inadequate quality and quantity was available for the determination of *MEF2C* in 99; *BAALC, HHEX, LYL1, TAL1, FAT1, XIST* and *TAL1* in 87 cases; *LMO2, DOT1L* and *LYN* in 78; LINC00648, *PCAT18* and LINC00461 in 72; *MEF2C-AS1, PCAT14* and LINC00202 in 76; *ST20, RAG1, EP300* and *EML4* in 84; *EZH2* and *KDM6A* in 82 and *MALAT1* in 81 cases due to inadequacy of the samples.

#### Association of protein and non-coding RNAs levels with patient variables

On analysis of the potential association of patient’s characteristics with expression levels of protein and non-coding RNAs, we found an association between *RAG1* expression and age of the patients. *RAG1* expression was higher in patients <12 years as compared to age ≥12 years (*p=*0.034). *XIST* and *KDM6A* expression were higher in females (*p=*0.047; 0.011, respectively). We did not find any association between WBC count at diagnosis and all parameters tested. Patients with low *XIST* expression and high *TAL1* more frequently had NCI high risk (*p=*0.01). Prednisolone resistance was associated with high *MEF2C* expression (*p=*0.048). Post-induction MRD positivity (≥0.01%) was associated with high expression of PCAT18 (*p=*0.04), *HHEX* (*p=*0.027) and *MEF2C* (*p=*0.007). (Table 2)

**Table 2:**
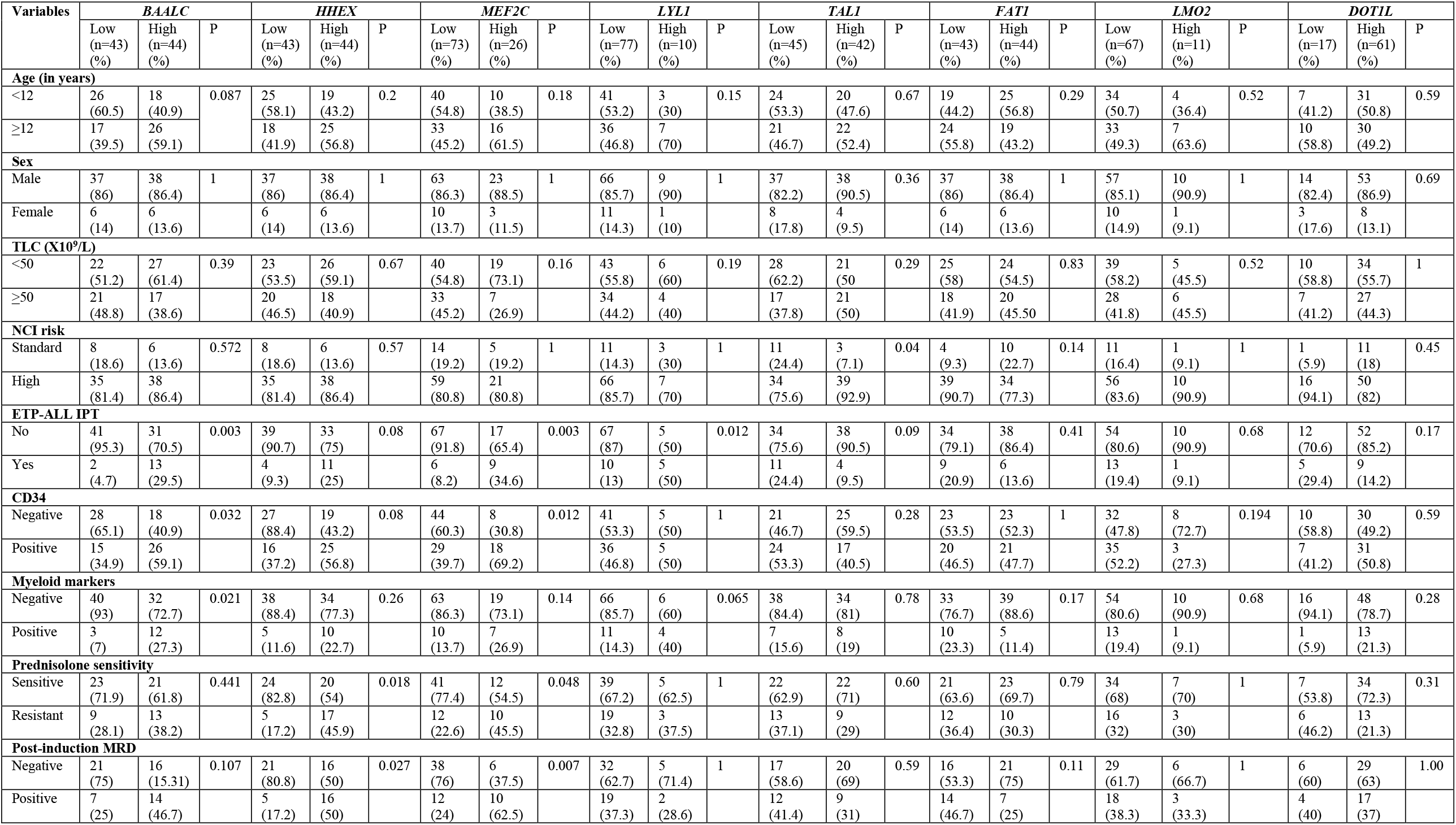
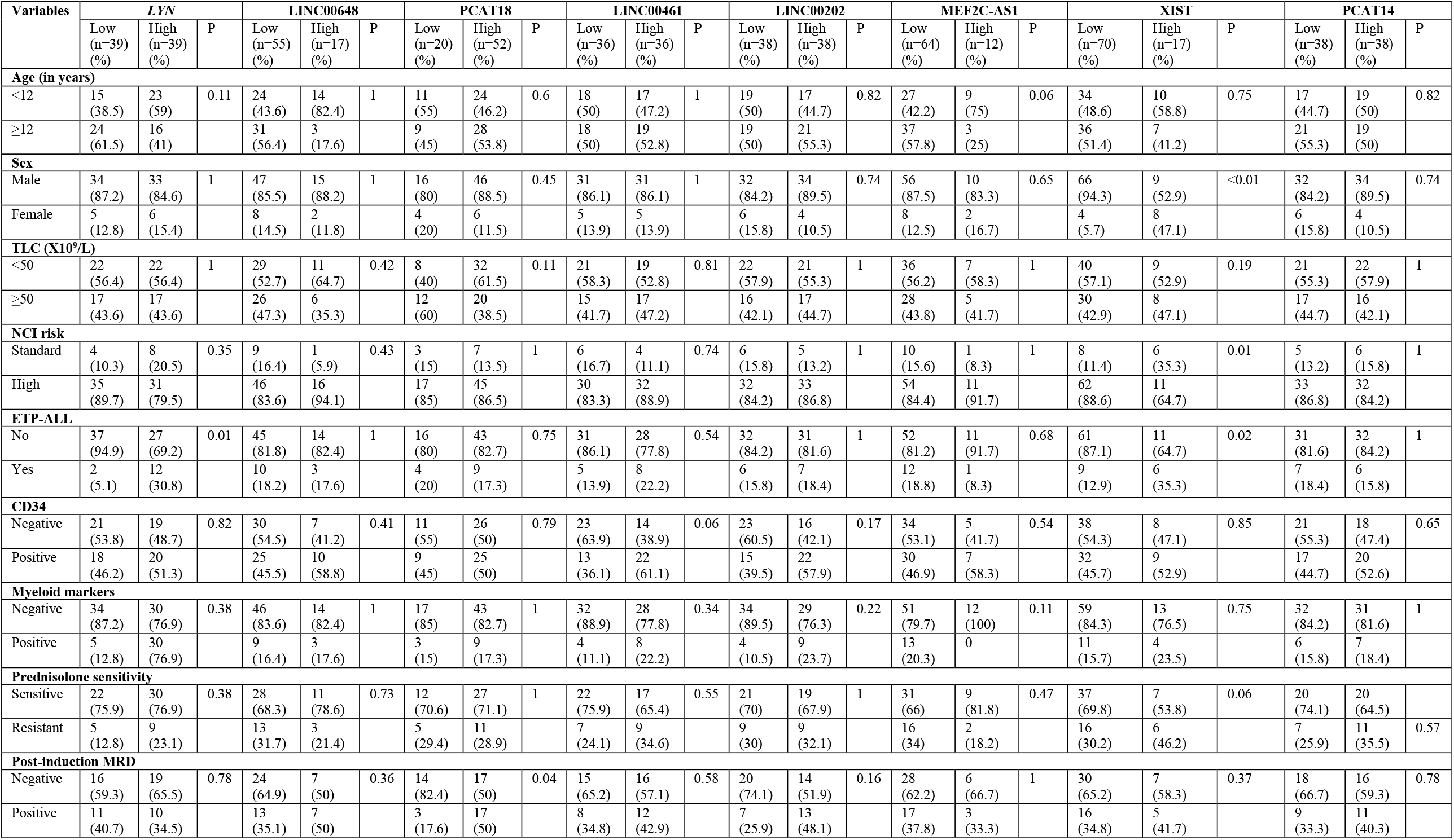
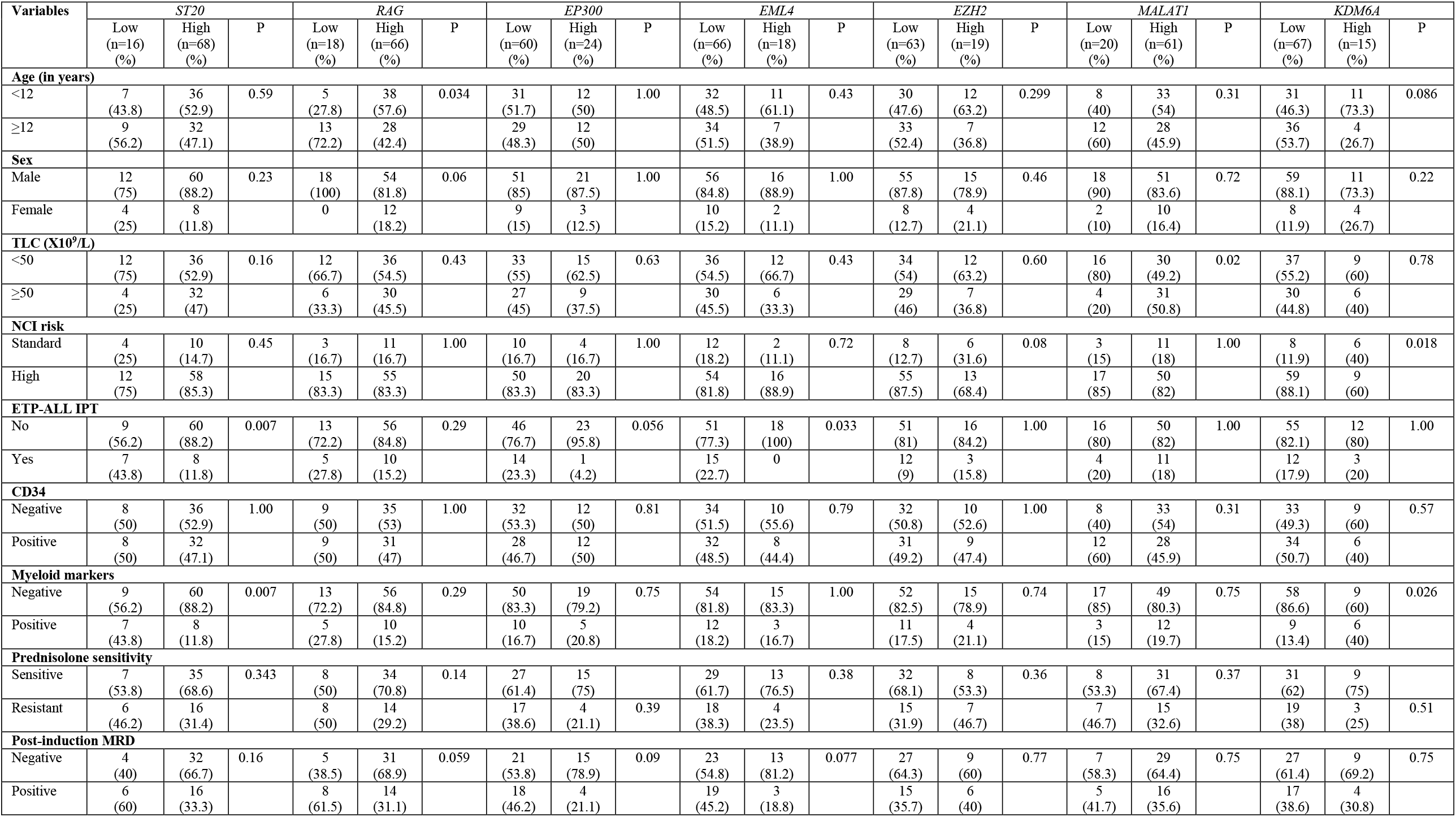
Association of expression of various protein and non-protein coding RNAs with patients’ characteristics.

### Survival analysis

Complete remission was achieved in 78 (87.64%) patients with induction chemotherapy. Median follow up was 22 months. The 3 year EFS (±SE) and OS (±SE) was 62.23±5.86% and 90.40±3.24%, respectively. On univariate analysis, we observed high *MEF2C* expression (low 71.78±6.58% vs high 36.57±10.74, HR 3.5, 95% confidence interval 1.67-7.3, *p=*0.0003), high *LYL1* expression (low 63.14±6.65% vs 26.67±15.9%, HR 2.69, 95% confidence interval 1.08-6.69, *p=*0.029), low *ST20* (low 43.21±13.56 vs high 61.02±7.26, HR 0.45, 95% confidence interval 0.19-1.03, *p=*0.049), low *RAG1* expression (low 41.67±12.9 vs high 61.24±7.49, HR 0.45, 95% confidence interval 0.20-0.99, *p=*0.037), low *EML4* expression (low 45.29±8.92 vs 83.33±8.78, HR 0.26, 95% confidence interval 0.078-0.88, *p=*0.018) and low *KDM6A* expression (low 50.48±7.58 vs high 83.33±8.78, HR 0.27, 95% confidence interval 0.062-1.12, *p=*0.049) were significantly associated with poor 3-year EFS (Table 3). In addition, age≥12 years and ETP-ALL immunophenotype were also associated with poor 3-year EFS (Table 3). We also found high *MEF2C* expression (low 94.77±2.96 vs high 78.75±8.45, HR 4.88, 95% confidence interval 1.16-20.40, *p=*0.016), low *DOT1L* expression (low 68.38±3.15 vs high 92.64±3.55, HR 0.22, 95% confidence interval 0.06-0.88, *p=*0.019), low *RAG1* expression (low 75±10.83 vs high 92.54±3.6, HR 0.25, 95% confidence interval 0.06-0.98, *p=*0.03) and low MALAT1 expression (low 76.02±10.48 vs high 94.11±3.31, HR 0.22, 95% confidence interval 0.048-0.97, *p=*0.027) to be significantly associated with poor 3-year OS (Table 3). On multivariate analysis for EFS, we found high *MEF2C* expression (HR 3.25, *p=*0.017) to be significantly associated with inferior EFS (Table 4a). We also found *MEF2C* expression (HR 6.73, *p=*0.04) and low MALAT1 expression (HR 0.16, *p=*0.031) to be significantly associated with inferior OS (Table 4b).

**Table 3:**
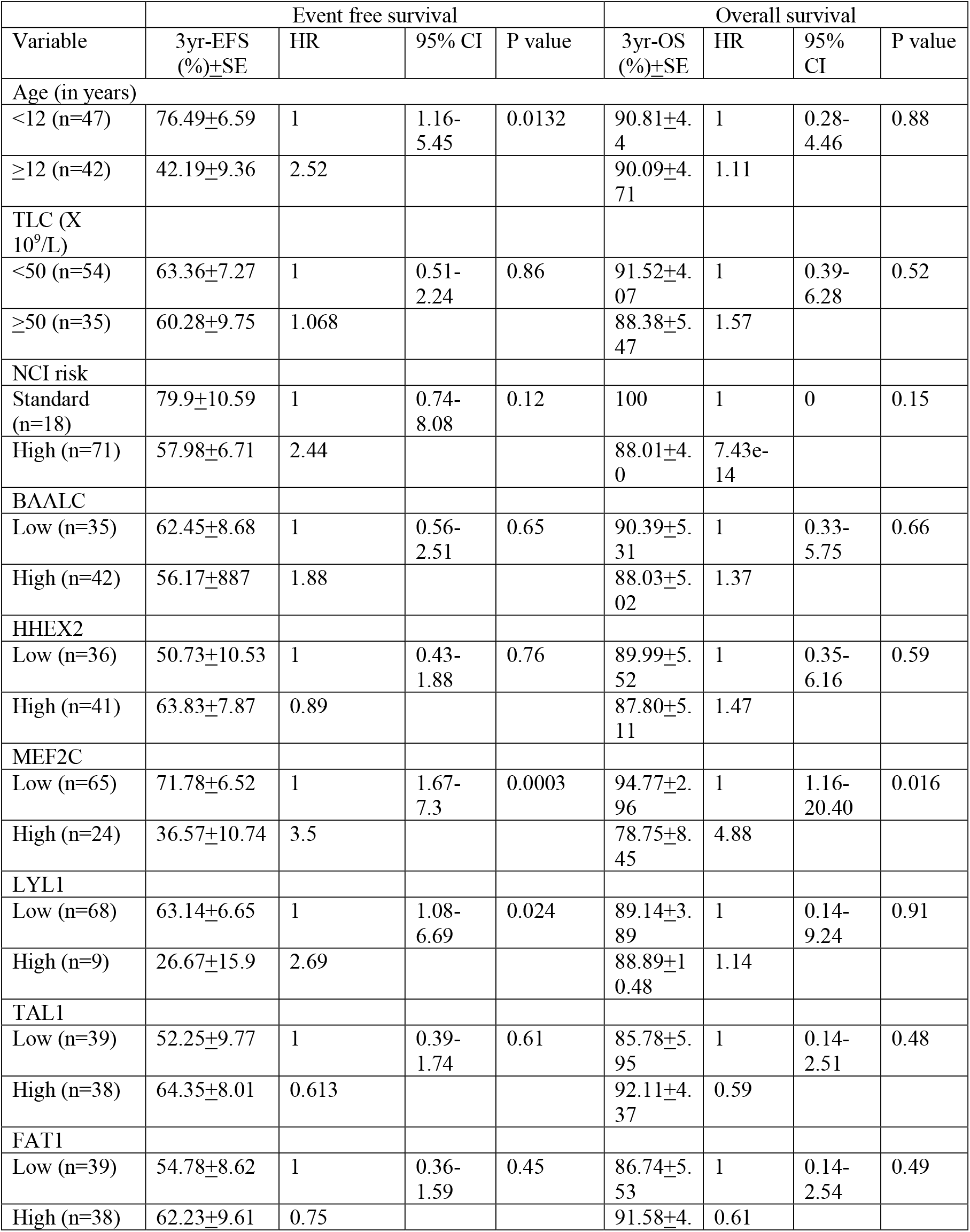
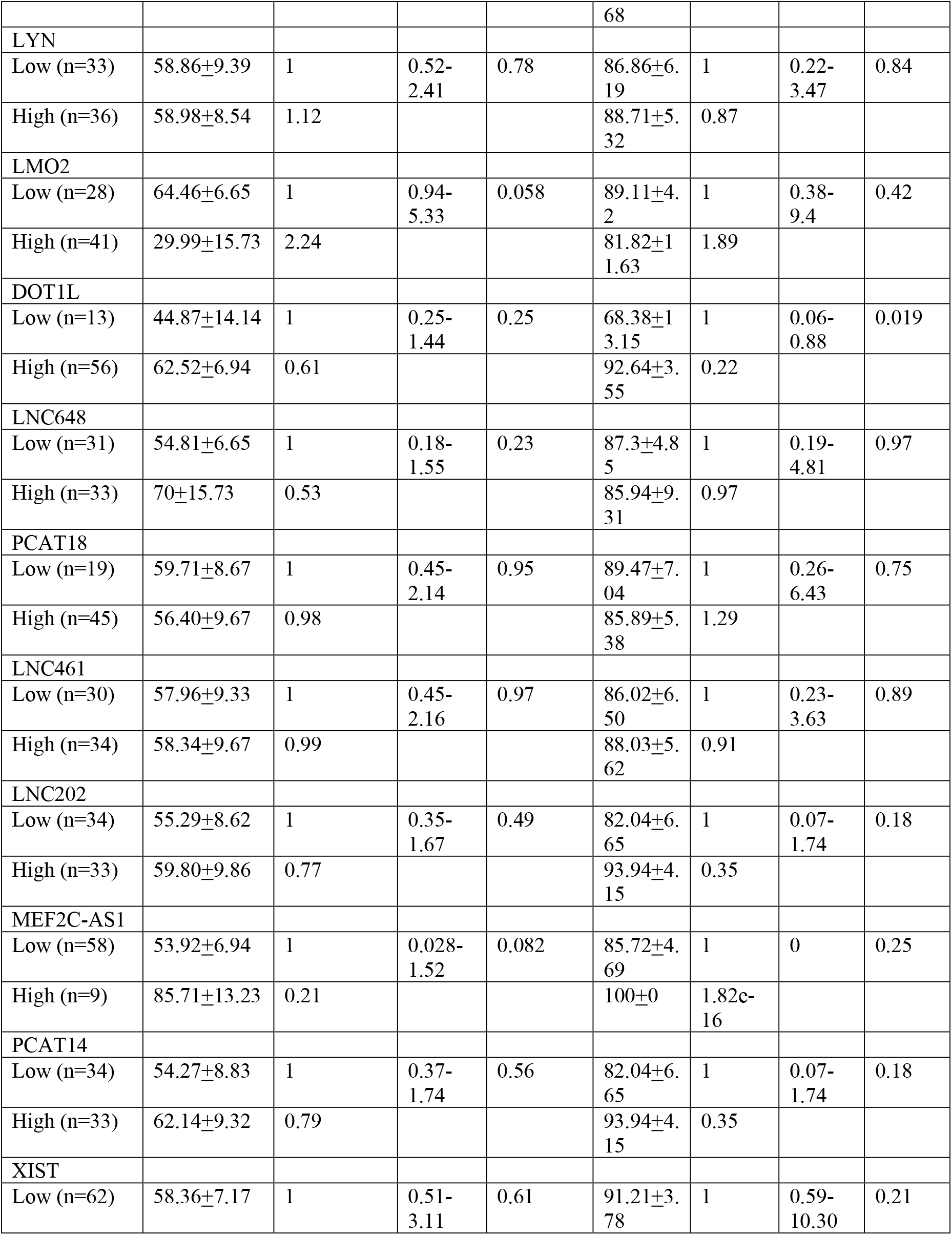
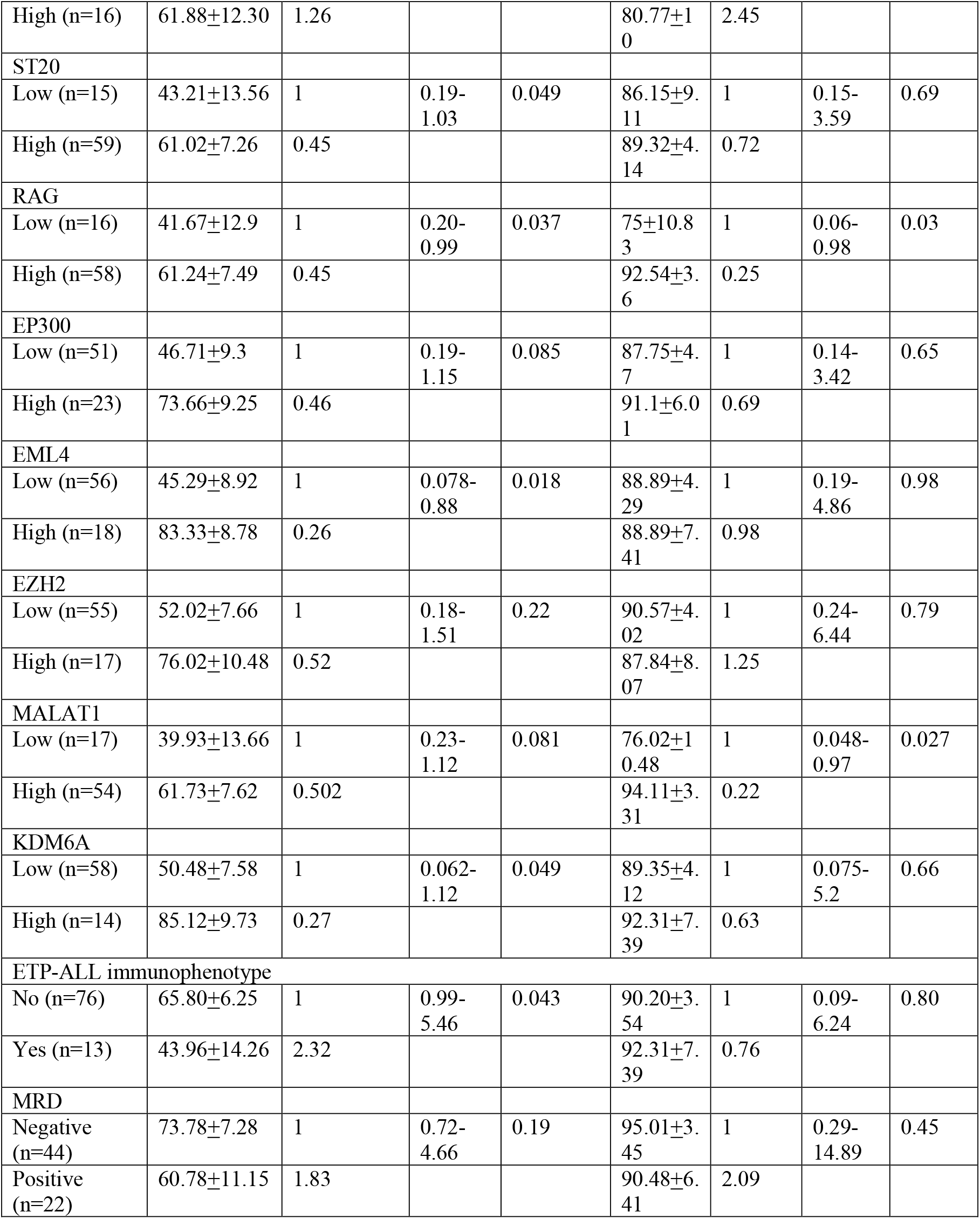
Association of patients’ characteristics with EFS and OS.

**Table 4:**
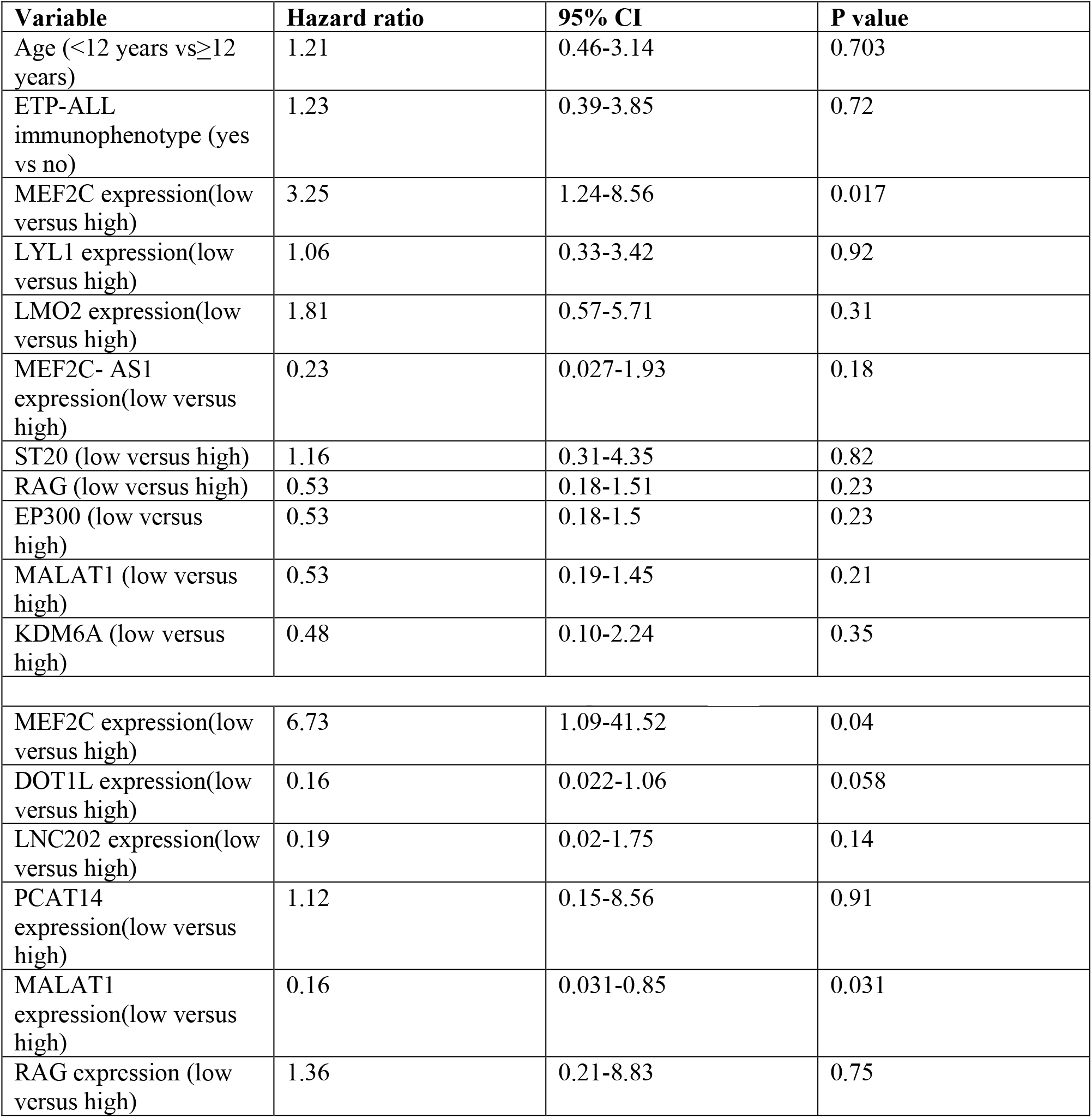
(a) Multivariate Cox regression analysis for EFS (b) Multivariate Cox regression analysis of OS.

## Discussion

With the advancement of molecular techniques, T-ALL has been extensively molecularly characterized. Although genetically heterogeneous, T-ALL can be categorized into various subtypes based on gene expression profiles. Although, unlike B-ALL, molecular features in T-ALL have not been utilized for risk stratification in clinical practice. Furthermore, only a limited number of studies have recently reported prognostic relevance of lncRNAs and epigenetic modifiers in T-ALL. In this study on 134 T-ALL cases, we performed high throughput RNA sequencing in the discovery cohort (n=35) and identified several protein-coding and non-coding transcripts which exhibit differential expression among immunophenotypic subtypes of T-ALL viz. immature, cortical and mature T-ALL. Furthermore, we also validated the expression of 23 identified targets in T-ALL patients and assessed their clinical significance.

The key genes which served as transcription factors in early hematopoiesis like *MEF2C, LYL1, LMO2, HHEX, RUNX2, HOXA10, HOXA9, RUNX1T1* and *ZBTB16* were upregulated in immature T-ALL.

*MEF2C* dysregulation has been previously shown in immature T-ALL^21-27^. Our previous study also showed the clinical importance of MEF2C in predicting prognosis of the ETP-ALL T-ALL patient (Singh et al 2020). Colomer-Lahiguera S, et al^22^, reported that *MEF2C* dysregulation in T-ALL is associated with CDKN1B deletion and poor response to prednisolone therapy. We also found an association between prednisolone resistance and high *MEF2C* expression. Starza et al^28^ showed its upregulation in T-ALL cases with interstitial deletion of 5q. Nagel et al^27^ proposed distinct mechanisms for aberrant *MEF2C* gene expression, either by NKX2-5 signaling or by chromosomal deletion of 5q. They also showed that *MEF2C* inhibits BCL2-regulated apoptosis by inhibition of NR4A1/NUR77^27^. In addition to this, Kawashima-Goto et al, 2015^23^, reported that BCL*2* inhibitors may be useful for treating T-ALL with high expression levels of *MEF2C*. On network analysis, *MEF2C* gene expression was found to interact with protein and non-protein-coding partners: *HOPX, KIT, BAALC, HHEX, EMP1, LYL1, BCL, SMYD3, HDAC9, HOTTIP, HOTAIR, LINC01021, XIST, MIR3142HG, MIR3132, MIR4741, SNORD100, SNORD101*.

In our study, *MN1, BAALC* and *IGFBP7* were overexpressed in immature T-ALL. The upregulation of these genes is believed to arise from T-cell progenitors retaining myeloid differentiation potential^29-31^. Previous studies suggest that overexpression of these genes is associated with poor outcome and resistance to chemotherapy^29,32-35^. Baldus et al, 2007^33^, reported that high *BAALC* expression was associated with poor long-term survival in T-ALL. On contrary to their observation, we did not find any significant association between *BAALC* expression and prednisolone sensitivity. Like previous studies, we found *BAALC* overexpression to be associated with the expression of CD34 and myeloid markers^29,31^.

However, we did not find any association between *BAALC* expression and patient outcome. We also found overexpression of *ZBTB16 (PLZF)* in our patients of immature T-ALL, although not stressed in previous western studies, was a notable finding in a recently reported study^21,36^. *ZBTB16* (or promyelocytic leukemia zinc finger, PLZF) contains one BTB domain and nine zinc finger motifs. Its overexpression was shown in this study to be a result of *ZBTB16-ABL1* translocation and occurred in different patients along with other mutations, including *NOTCH1, ZEB2, PTEN, MYCN*, and *PIK3CD*. Laboratory studies with both *in vitro* and mouse model suggest *ZBTB16-ABL1* be a driver leukemogenic lesion that causes increased proliferation and a heightened protein tyrosine kinase (PTK) activity that is amenable to tyrosine kinase inhibitor (TKI) activity^21^. These findings indicate that our *ZBTB16*-expressing patients have the same translocation. Our finding is of significance also because along with *LYN* overexpression, T-ALL patients with ZBTB16 overexpression may also benefit from TKIs.

Apart from these known genes, we identified aberrantly expression of some genes which have not been reported in T-ALL patients such as *RUNX1T1, RUNX2, PLD4, NT5E (CD73), HOPX, TP63, HOXA11-AS*. A role for *RUNX2* in T-ALL has been suggested in a study by Nagel et al, 2011, who, to uncover additional target genes, investigated in detail the aberrant expression of *MEF2C* mediated by complex deletion at 5q, del(5)(q14) in T-ALL cell line^26^. This could be an evidential proof where RUNX2 instead of RUNX1 could be involved in the manifestation of ETP-ALL that allows *in vivo* functional evaluation of putative oncogenes and allows preclinical drug testing.

Further, some of the observed differentially expressed gene in cortical T-ALL such as *CD1A, CD1C, CD4, CFTR, FAT3, NKX2-1, TLX1, TLX3* and *RAG1* have been previously reported while we observed three additional genes, *EREG, PAX* and, *ZIC2* to be upregulated in the present study.

Neumann et al 2013, in a study of adult ETP-ALL, showed that cadherins *FAT1* (25%) and *FAT3* (20%) were mutated, implicating alterations in cell adhesion, and activation of the Wnt pathway^37^. Neumann et al, 2014^38^, showed that *FAT1* expression was correlated with a more mature leukemic immunophenotype in T-ALL, with 74% patients with thymic T-ALL being *FAT1* positive compared with 45% of patients with mature T-ALL and only 4% of early T-ALL patients. This is in line with our results, as we observed that FAT1 was associated with cortical immunophenotype in our study. Like previous study^38^, we did not find any correlation between FAT1 expression and patient outcome.

Mature T-ALL is a rare subgroup and immunophenotypically diagnosed by CD1a- and sCD3+. Molecularly *TAL1* has been identified as a driver gene for late cortical T-ALL^1^. We observed *TAL1* be overexpressed in both mature and cortical T-ALL in our study. Among the protein-coding genes, *APC2, BCL3, CCR4, ST20, EML4* and *NCOR2* were some of the key upregulated genes.

Aberrant histone modifications are the hallmark for cancer and are associated with dysregulated expression of histone modifiers. Therefore, we also studied their expression pattern of epigenetic modifiers to identify a set of histones modifying enzymes to be upregulated or downregulated specifically to the subtype of T-ALL. *EZH2*, a member of the polycomb repressor complex, which is known to be under-expressed in our immature T-ALL cases. This may be related to their previously reported mutations in immature T-ALL^39^. Danis et al^40^, mechanistically linked *EZH2* inactivation to stem-cell-associated transcriptional programs and increased growth/survival signaling, features that convey an adverse prognosis in patients. However, we did not observe the association between *EZH2* expression and clinical outcome. Loss-of-function mutations and deletions in *SETD2* have been shown to lead to chemotherapy tolerance and clonal survival by cell cycle arrest followed by apoptosis. Further, overexpression of *SETD2* has been demonstrated to confer chemotherapy resistance in a variety of cancers including leukemias^39,41-43^. We also observed overexpression of *SETD2* in T-ALL as compared to the normal thymus. Therefore, the potential role of SETD2 overexpression in therapeutic resistance in T-ALL requires further investigation. In pediatric cases, higher expression of *HDAC7* and *HDAC9* in ALL is associated with poor prognosis. In our study, we observed overexpression of *HDAC9* in our immature and cortical cases. *CREBBP, EP300, ASH1L, ATM, PKN1, KDM2B, KDM4B* and *DOT1L* showed significant differential expression in mature T-ALL. In the context of transcription coactivation, *EP300* and *CREBBP* have lysine acetyltransferase activity^44-47^. Targeted histone lysine acetylation of *EP300* and *CREBBP* can influence chromatin conformation^46^, and concomitant binding of *EP300* and acetylation of H3K27 is a hallmark of promoter or enhancer activation^48^.

We also found low expression of *ST20* (figure 5c)and *EML4*(figure 5e) to be associated with poor EFS. This has not been reported before.

**Figure 5.**
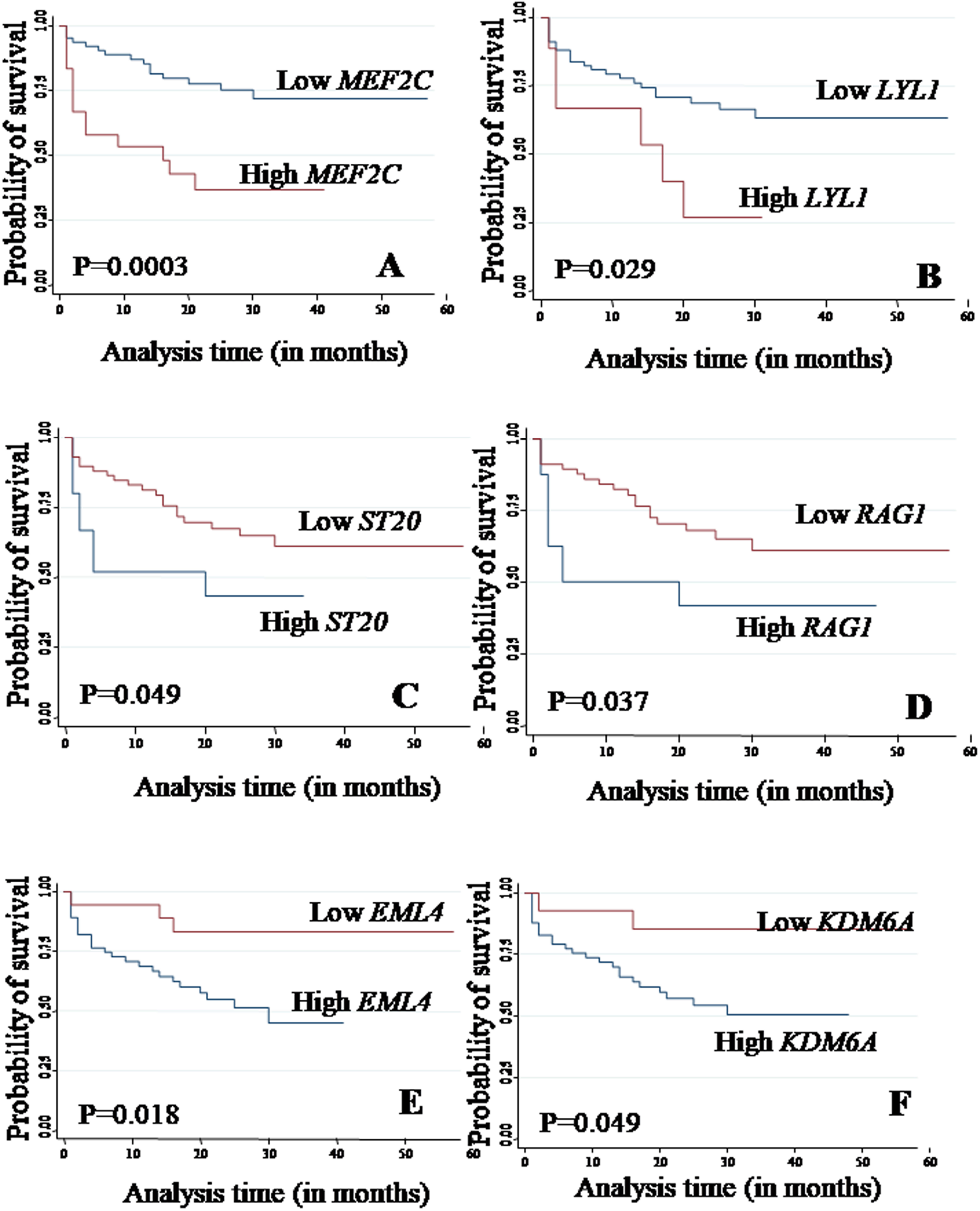
Kaplan Meier analysis for event free survival for expression of **a**. *MEF2C*, **b**. *LYL1*, **c**. *ST20*, **d**. *RAG1*, **e**. *EML4* and **f**. *KDM6A* genes.

DOT1-like (*DOT1L*) histone lysine methyltransferase methylates H3K79 and plays an important role in embryogenesis and hematopoiesis (Ref). Its aberrant activation is associated with acute leukemias^49,50^, but Its function is unknown in T-ALL. *DOT1L* catalytic activity depends on the mono-ubiquitination of lysine120 in histone H2B (H2BK120Ub), which provides crosstalk between various histone post-translational modifications^51^. Recent studies suggested the role of *DOT1L* in H3K79 methylation and mono-ubiquitination of lysine (H2BK120Ub) that may pave the way for the development of novel *DOT1L*-driven anti-leukemia therapies.^52-56^. *DOT1L* was overexpressed in our mature T-ALL patients and it may be worth investigating if they could be subjects for *DOT1L*-driven anti-leukemia therapy. We found *DOT1L* low expression to be associated with poor OS. This has never been reported before.

Apart from proteins, non-coding transcript’s/ RNA repertoire forms another layer of regulatory paradigm in normal cell hemostasis. Using RNA-seq, we identified the differentially expressed non-coding RNAs especially lncRNAs which have been very well documented earlier for their role in cancers. Our analysis revealed 223 lncRNAs, showing differential expression among various T-ALL subtypes. *NOTCH1*-regulated lncRNA, LUNAR1, was overexpressed in cortical and mature T-ALL^20^. This may be related to a higher incidence of activating *NOTCH1*1 mutations in this T-ALL subtypes^1^. HOTTIP and *MEF2C*-AS1 were overexpressed in immature; LINC00202, LINC0648, LINC00461 in cortical T-ALL and MALAT1 in mature T-ALL. These have not been reported in T-ALL before in the English literature. We did not find any significant association between their expression with immunophenotypic subtypes.

HOTTIP has been reported to be aberrantly activated in AML. It promotes hematopoietic stem cell renewal leading to AML-like disease in mice.^57^ This may explain its overexpression in immature T-ALL which has myeloid potential in our study. MALAT1 is known to be involved in a plethora of biological processes ranging from alternative splicing, nuclear organization, epigenetic regulation of gene expression. It is also associated with various pathological complications like breast cancer, lung adenocarcinomas, hepatocellular carcinomas, bladder cancers and diabetes etc^58-60^. Several studies suggest MALAT1 expression as a prognostic marker for various cancer types^61^. At a molecular level, MALAT1 plays an important role in modulating several signaling pathways like *MAPK/ERK, PI3K/AKT, WNT* and *NF-kB* leading to a modification of proliferation, cell death, cell cycle, migration, invasion, immunity, angiogenesis, and tumorigenicity. The exact mechanism of how MALAT1 helps in cancer development and progression is not fully known. MALAT1 can be a therapeutic target, potential diagnostic and prognosis biomarker for cancers^59,62,63^.

Out of the 23 targets tested, we found that *MEF2C* gene expression emerged as a significant predictor of EFS and OS(figure 5a ans 6a). Although *MEF2C* overexpression is associated with chemoresistance and poor outcome in AML, its prognostic relevance in T-ALL has not been reported to the best of our knowledge. MALAT1 low expression also emerged as a marker for the poor OS. This has also not been reported before. Apart from these, the prognostic relevance of other markers like, *ST20*(figure 5c) *DOT1L, RAG1*(figure 5d), *EML4* (figure 5e) *and KDM6a* (figure 5f) should be studied in a larger number of patients. Previous studies on the utilization of high throughput sequencing and microarray gene expression have shown that the immature gene signature is associated with inferior survival in T-ALL. Both of these methods are time, labour and cost-intensive. Based on our results, we recommend *MEF2C* gene expression analysis by real-time PCR is a reliable and cheap alternative, therefore, can be easily integrated into routine clinical practice.

**Figure 6.**
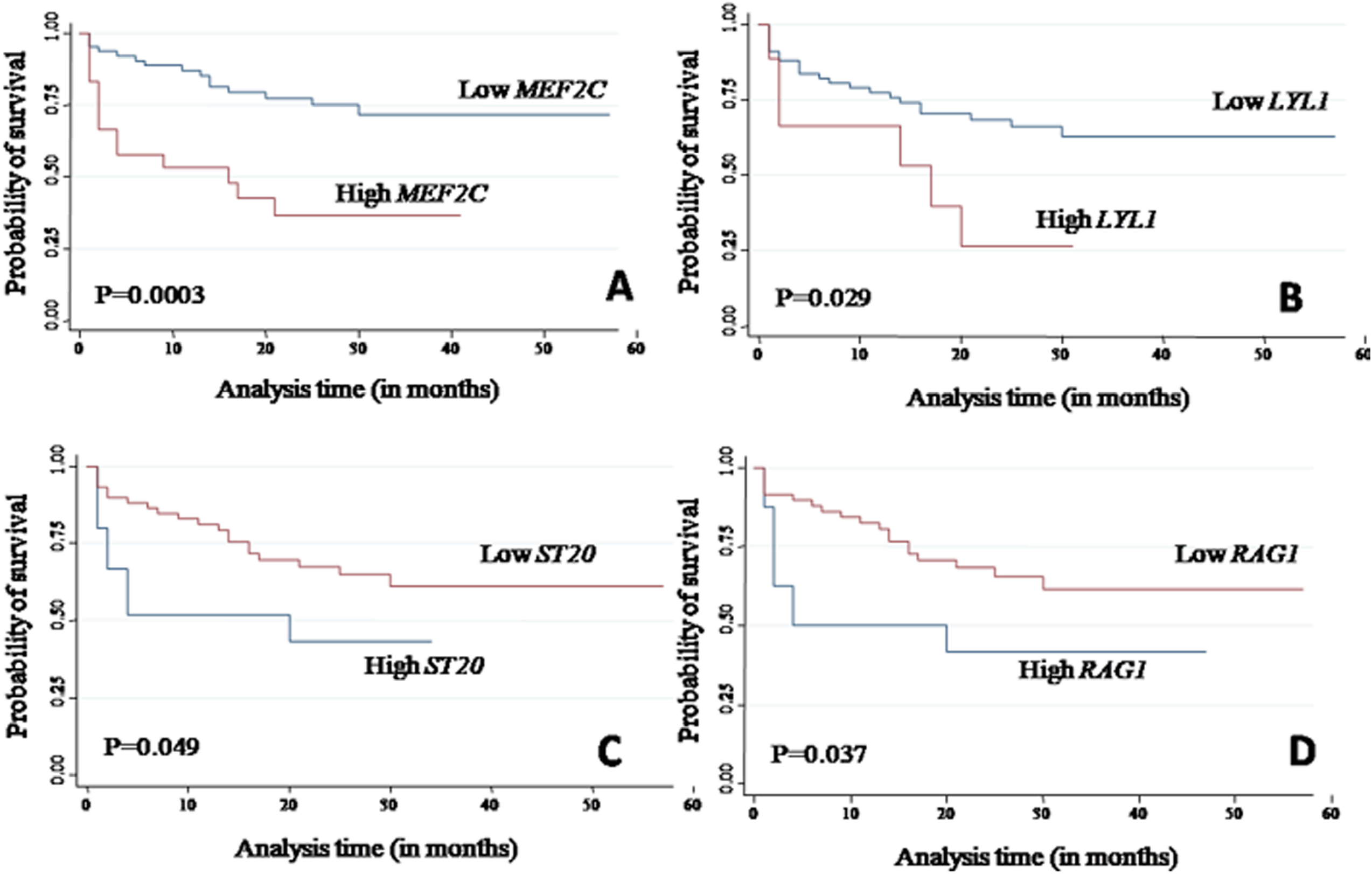
Kaplan Meier analysis for overall survival for expression of **a**. *MEF2C*, **b**. *DOT1L*, **c**. *RAG1*, **d**. *MALAT1*.

Taken together, our study provides a comprehensive transcriptional map of coding as well as long noncoding RNAs. We have identified unique gene signatures that were not discovered in the western population. This may be related to the ethnic variation of Indian patients. Along with protein-coding genes, we identified novel as well as known lncRNAs which were differentially expressed in T-ALL patients. Experimental validation and survival analysis for some of the candidates confirmed the RNA-seq results while co-expression analysis gave an insight into the putative functional roles and pathways involved in T-ALL. *MEF2C* high expression emerged as a significant predictor of poor EFS and OS.

## Data Availability

Data will be available on kind request after publication.

## Acknowledgements

None

## Ethics approval and consent to participate

I confirm all relevant ethical guidelines have been followed, and necessary ethical approval approvals (IESC/T-395/28-11-2014) have been obtained from Institute Ethics Committee For Post Graduate Research, All India Institute of Medical Sciences New Delhi.

## Author Contribution

DV performed the wetlab experiments related to RNA-Seq and also analyzed the data with SKp, DS. JS, GS, SKr, MA performed the validation experiments and data collection. SB, RS, BN, AS, RP, JP, RK contributed to project design, clinical data, patient recruitment, and experiment management. SS,VS supported through highthroughput computing facility. AC is the principal project investigator.

## References

1. Belver, L. and A. Ferrando (2016). “The genetics and mechanisms of T cell acute lymphoblastic leukaemia.” Nature Reviews Cancer 16: 494.

2. Van Vlierberghe, P., A. Ambesi-Impiombato, K. De Keersmaecker, M. Hadler, E. Paietta, M. S. Tallman, J. M. Rowe, C. Forne, M. Rue and A. A. Ferrando (2013). “Prognostic relevance of integrated genetic profiling in adult T-cell acute lymphoblastic leukemia.” Blood 122(1): 74.

3. Gianni F, Belver L, Ferrando A. The Genetics and Mechanisms of T-Cell Acute Lymphoblastic Leukemia. Cold Spring Harb Perspect Med. 2020 Mar 2;10(3). pii:a035246.

4. Van Vlierberghe, P. and A. Ferrando (2012). “The molecular basis of T cell acute lymphoblastic leukemia.” J Clin Invest 122(10): 3398–3406.

5. Correia, N. C., A. Girio, I. Antunes, L. R. Martins and J. T. Barata (2014). “The multiple layers of non-genetic regulation of PTEN tumour suppressor activity.” Eur J Cancer 50(1): 216–225.

6. Fragoso, R. and J. T. Barata (2014). “PTEN and leukemia stem cells.” Advances in Biological Regulation 56: 22–29.

7. Fragoso, R. and J. T. Barata (2015). “Kinases, tails and more: Regulation of PTEN function by phosphorylation.” Methods 77-78: 75–81.

8. Martelli, A. M., G. Tabellini, F. Ricci, C. Evangelisti, F. Chiarini, R. Bortul, J. A. McCubrey and F. A. Manzoli (2012). “PI3K/AKT/mTORC1 and MEK/ERK signaling in T-cell acute lymphoblastic leukemia: New options for targeted therapy.” Advances in Biological Regulation 52(1): 214–227.

9. Girardi T, Vicente C, Cools J, De Keersmaecker K. The genetics and molecular biology of T-ALL. Blood. 2017 Mar 2;129(9):1113–1123.

10. Iacobucci I, Mullighan CG. Genetic Basis of Acute Lymphoblastic Leukemia. J Clin Oncol. 2017 Mar 20;35(9):975–983.

11. Taylor J, Xiao W, Abdel-Wahab O. Diagnosis and classification of hematologic malignancies on the basis of genetics. Blood. 2017 Jul 27;130(4):410–423.

12. Coustan-Smith, E., C. G. Mullighan, M. Onciu, F. G. Behm, S. C. Raimondi, D. Pei, C. Cheng, X. Su, J. E. Rubnitz, G. Basso, A. Biondi, C.-H. Pui, J. R. Downing and D. Campana (2009). “Early T-cell precursor leukaemia: a subtype of very high-risk acute lymphoblastic leukaemia.” The Lancet Oncology 10(2): 147–156.

13. Bene MC, Castoldi G, Knapp W, Ludwig WD, Matutes E, Orfao A, van’t Veer MB. Proposals for the immunological classification of acute leukemias. European Group for the Immunological Characterization of Leukemias (EGIL). Leukemia. 1995 Oct;9(10):1783–6.

14. Huang da, W., B. T. Sherman and R. A. Lempicki (2009). “Bioinformatics enrichment tools: paths toward the comprehensive functional analysis of large gene lists.” Nucleic Acids Res 37(1): 1–13.

15. Huang da, W., B. T. Sherman and R. A. Lempicki (2009). “Systematic and integrative analysis of large gene lists using DAVID bioinformatics resources.” Nat Protoc 4(1): 44–57.

16. Shannon, P., A. Markiel, O. Ozier, N. S. Baliga, J. T. Wang, D. Ramage, N. Amin, B. Schwikowski and T. Ideker (2003). “Cytoscape: a software environment for integrated models of biomolecular interaction networks.” Genome Res 13(11): 2498–2504.

17. Haferlach C, Kern W, Schindela S, Kohlmann A, Alpermann T, Schnittger S,Haferlach T. Gene expression of BAALC, CDKN1B, ERG, and MN1 adds independent prognostic information to cytogenetics and molecular mutations in adult acute myeloid leukemia. Genes Chromosomes Cancer. 2012 Mar;51(3):257–65.

18. Heesch, S., C. Schlee, M. Neumann, A. Stroux, A. Kuhnl, S. Schwartz, T. Haferlach, N. Goekbuget, D. Hoelzer, E. Thiel, W. K. Hofmann and C. D. Baldus (2010). “BAALC-associated gene expression profiles define IGFBP7 as a novel molecular marker in acute leukemia.” Leukemia 24(8): 1429–1436.

19. Yin S, Dou J, Yang G, Chen F. Long non-coding RNA XIST expression as a prognostic factor in human cancers: A meta-analysis. Int J Biol Markers. 2019 Dec;34(4):327–333.

20. Trimarchi T, Bilal E, Ntziachristos P, Fabbri G, Dalla-Favera R, Tsirigos A,Aifantis I. Genome-wide mapping and characterization of Notch-regulated long noncoding RNAs in acute leukemia. Cell. 2014 Jul 31;158(3):593–606.

21. Chen B, Jiang L, Zhong ML, Li JF, Li BS, Peng LJ, Dai YT, Cui BW, Yan TQ,Zhang WN, Weng XQ, Xie YY, Lu J, Ren RB, Chen SN, Hu JD, Wu DP, Chen Z, Tang JY,Huang JY, Mi JQ, Chen SJ. Identification of fusion genes and characterization of transcriptome features in T-cell acute ymphoblastic leukemia. Proc Natl Acad Sci U S A. 2018 Jan 9;115(2):373–378.

22. Colomer-Lahiguera S, Pisecker M, König M, Nebral K, Pickl WF, Kauer MO, Haas OA, Ullmann R, Attarbaschi A, Dworzak MN, Strehl S. MEF2C-dysregulated pediatric T-cell acute lymphoblastic leukemia is associated with CDKN1B deletions and a poor response to glucocorticoid therapy. Leuk Lymphoma. 2017 Dec;58(12):2895–2904.

23. Kawashima-Goto S, Imamura T, Tomoyasu C, Yano M, Yoshida H, Fujiki A, Tamura S, Osone S, Ishida H, Morimoto A, Kuroda H, Hosoi H. BCL2 Inhibitor (ABT-737): A Restorer of Prednisolone Sensitivity in Early T-Cell Precursor-Acute Lymphoblastic Leukemia with High MEF2C Expression? PLoS One. 2015 Jul 14;10(7):e0132926.

24. Zuurbier L, Gutierrez A, Mullighan CG, Canté-Barrett K, Gevaert AO, de Rooi J,Li Y, Smits WK, Buijs-Gladdines JG, Sonneveld E, Look AT, Horstmann M, Pieters R,Meijerink JP. Immature MEF2C-dysregulated T-cell leukemia patients have an early T-cell precursor acute lymphoblastic leukemia gene signature and typically have non-rearranged T-cell receptors. Haematologica. 2014 Jan;99(1):94–102.

25. Homminga I, Pieters R, Langerak AW, de Rooi JJ, Stubbs A, Verstegen M, Vuerhard M, Buijs-Gladdines J, Kooi C, Klous P, van Vlierberghe P, Ferrando AA,Cayuela JM, Verhaaf B, Beverloo HB, Horstmann M, de Haas V, Wiekmeijer AS,Pike-Overzet K, Staal FJ, de Laat W, Soulier J, Sigaux F, Meijerink JP.Integrated transcript and genome analyses reveal NKX2-1 and MEF2C as potential oncogenes in T cell acute lymphoblastic leukemia. Cancer Cell. 2011 Apr 12;19(4):484–97.

26. Nagel S, Venturini L, Meyer C, Kaufmann M, Scherr M, Drexler HG, Macleod RA. Transcriptional deregulation of oncogenic myocyte enhancer factor 2C in T-cell acute lymphoblastic leukemia. Leuk Lymphoma. 2011 Feb;52(2):290–7.

27. Nagel S, Meyer C, Quentmeier H, Kaufmann M, Drexler HG, MacLeod RA. MEF2C is activated by multiple mechanisms in a subset of T-acute lymphoblastic leukemia cell lines. Leukemia. 2008 Mar;22(3):600–7.

28. La Starza R, Barba G, Demeyer S, Pierini V, Di Giacomo D, Gianfelici V, Schwab C, Matteucci C, Vicente C, Cools J, Messina M, Crescenzi B, Chiaretti S, Foà R,Basso G, Harrison CJ, Mecucci C. Deletions of the long arm of chromosome 5 define subgroups of T-cell acute lymphoblastic leukemia. Haematologica. 2016 Aug;101(8):951–8.

29. Heesch S, Bartram I, Neumann M, Reins J, Mossner M, Schlee C, Stroux A, Haferlach T, Goekbuget N, Hoelzer D, Hofmann WK, Thiel E, Baldus CD. Expression of IGFBP7 in acute leukemia is regulated by DNA methylation. Cancer Sci. 2011 Jan;102(1):253–9.

30. Neumann M, Heesch S, Gökbuget N, Schwartz S, Schlee C, Benlasfer O, Farhadi-Sartangi N, Thibaut J, Burmeister T, Hoelzer D, Hofmann WK, Thiel E,Baldus CD. Clinical and molecular characterization of early T-cell precursor leukemia: a high-risk subgroup in adult T-ALL with a high frequency of FLT3 mutations. Blood Cancer J. 2012 Jan;2(1):e55.

31. Heesch S, Schlee C, Neumann M, Stroux A, Kühnl A, Schwartz S, Haferlach T,Goekbuget N, Hoelzer D, Thiel E, Hofmann WK, Baldus CD. BAALC-associated gene expression profiles define IGFBP7 as a novel molecular marker in acute leukemia. Leukemia. 2010 Aug;24(8):1429–36.

32. Carow, C. E., M. Levenstein, S. H. Kaufmann, J. Chen, S. Amin, P. Rockwell, L. Witte, M. J. Borowitz, C. I. Civin and D. Small (1996). “Expression of the hematopoietic growth factor receptor FLT3 (STK-1/Flk2) in human leukemias.” Blood 87(3): 1089.

33. Baldus, C. D., P. Martus, T. Burmeister, S. Schwartz, N. Gokbuget, C. D. Bloomfield, D. Hoelzer, E. Thiel and W. K. Hofmann (2007). “Low ERG and BAALC expression identifies a new subgroup of adult acute T-lymphoblastic leukemia with a highly favorable outcome.” J Clin Oncol 25(24): 3739–3745.

34. Baldus, C. D., S. M. Tanner, D. F. Kusewitt, S. Liyanarachchi, C. Choi, M. A. Caligiuri, C. D. Bloomfield and A. d. l. Chapelle (2003). “BAALC, a novel marker of human hematopoietic progenitor cells.” Experimental Hematology 31(11): 1051–1056.

35. Heuser, M., B. Argiropoulos, F. Kuchenbauer, E. Yung, J. Piper, S. Fung, R. F. Schlenk, K. Dohner, T. Hinrichsen, C. Rudolph, A. Schambach, C. Baum, B. Schlegelberger, H. Dohner, A. Ganser and R. K. Humphries (2007). “MN1 overexpression induces acute myeloid leukemia in mice and predicts ATRA resistance in patients with AML.” Blood 110(5): 1639.

36. Ferrando, A. A., D. S. Neuberg, J. Staunton, M. L. Loh, C. Huard, S. C. Raimondi, F. G. Behm, C.-H. Pui, J. R. Downing, D. G. Gilliland, E. S. Lander, T. R. Golub and A. T. Look (2002). “Gene expression signatures define novel oncogenic pathways in T cell acute lymphoblastic leukemia.” Cancer Cell 1(1): 75–87.

37. Neumann, M., E. Coskun, L. Fransecky, L. H. Mochmann, I. Bartram, N. F. Sartangi, S. Heesch, N. Gokbuget, S. Schwartz, C. Brandts, C. Schlee, R. Haas, U. Duhrsen, M. Griesshammer, H. Dohner, G. Ehninger, T. Burmeister, O. Blau, E. Thiel, D. Hoelzer, W. K. Hofmann and C. D. Baldus (2013). “FLT3 mutations in early T-cell precursor ALL characterize a stem cell like leukemia and imply the clinical use of tyrosine kinase inhibitors.” PLoS One 8(1): e53190.

38. Neumann, M., M. Seehawer, C. Schlee, S. Vosberg, S. Heesch, E. K. von der Heide, A. Graf, S. Krebs, H. Blum, N. Gökbuget, S. Schwartz, D. Hoelzer, P. A. Greif and C. D. Baldus (2014). “FAT1 expression and mutations in adult acute lymphoblastic leukemia.” Blood Cancer Journal 4: e224.

39. Zhang, J., L. Ding, L. Holmfeldt, G. Wu, S. L. Heatley, D. Payne-Turner, J. Easton, X. Chen, J. Wang, M. Rusch, C. Lu, S. C. Chen, L. Wei, J. R. Collins-Underwood, J. Ma, K. G. Roberts, S. B. Pounds, A. Ulyanov, J. Becksfort, P. Gupta, R. Huether, R. W. Kriwacki, M. Parker, D. J. McGoldrick, D. Zhao, D. Alford, S. Espy, K. C. Bobba, G. Song, D. Pei, C. Cheng, S. Roberts, M. I. Barbato, D. Campana, E. Coustan-Smith, S. A. Shurtleff, S. C. Raimondi, M. Kleppe, J. Cools, K. A. Shimano, M. L. Hermiston, S. Doulatov, K. Eppert, E. Laurenti, F. Notta, J. E. Dick, G. Basso, S. P. Hunger, M. L. Loh, M. Devidas, B. Wood, S. Winter, K. P. Dunsmore, R. S. Fulton, L. L. Fulton, X. Hong, C. C. Harris, D. J. Dooling, K. Ochoa, K. J. Johnson, J. C. Obenauer, W. E. Evans, C. H. Pui, C. W. Naeve, T. J. Ley, E. R. Mardis, R. K. Wilson, J. R. Downing and C. G. Mullighan (2012). “The genetic basis of early T-cell precursor acute lymphoblastic leukaemia.” Nature 481(7380): 157–163.

40. Danis E, Yamauchi T, Echanique K, Zhang X, Haladyna JN, Riedel SS, Zhu N, Xie H, Orkin SH, Armstrong SA, Bernt KM, Neff T. EZH2 Controls an Early Hematopoietic Program and Growth and Survival Signaling in Early T Cell Precursor Acute Lymphoblastic Leukemia. Cell Rep. 2016 Mar 1;14(8):1953–65.

41. Mar, B. G., L. B. Bullinger, K. M. McLean, P. V. Grauman, M. H. Harris, K. Stevenson, D. S. Neuberg, A. U. Sinha, S. E. Sallan, L. B. Silverman, A. L. Kung, L. Lo Nigro, B. L. Ebert and S. A. Armstrong (2014). “Mutations in epigenetic regulators including SETD2 are gained during relapse in paediatric acute lymphoblastic leukaemia.” Nat Commun 5: 3469.

42. Zhu, X., F. He, H. Zeng, S. Ling, A. Chen, Y. Wang, X. Yan, W. Wei, Y. Pang, H. Cheng, C. Hua, Y. Zhang, X. Yang, X. Lu, L. Cao, L. Hao, L. Dong, W. Zou, J. Wu, X. Li, S. Zheng, J. Yan, J. Zhou, L. Zhang, S. Mi, X. Wang, L. Zhang, Y. Zou, Y. Chen, Z. Geng, J. Wang, J. Zhou, X. Liu, J. Wang, W. Yuan, G. Huang, T. Cheng and Q.-f. Wang (2014). “Identification of functional cooperative mutations of SETD2 in human acute leukemia.” Nature Genetics 46: 287.

43. Wang, S., X. Yuan, Y. Liu, K. Zhu, P. Chen, H. Yan, D. Zhang, X. Li, H. Zeng, X. Zhao, X. Chen, G. Zhou and S. Cao (2019). “Genetic polymorphisms of histone methyltransferase SETD2 predicts prognosis and chemotherapy response in Chinese acute myeloid leukemia patients.” J Transl Med 17(1): 101.

44. Ait-Si-Ali, S., S. Ramirez, F. X. Barre, F. Dkhissi, L. Magnaghi-Jaulin, J. A. Girault, P. Robin, M. Knibiehler, L. L. Pritchard, B. Ducommun, D. Trouche and A. Harel-Bellan (1998). “Histone acetyltransferase activity of CBP is controlled by cycle-dependent kinases and oncoprotein E1A.” Nature 396(6707): 184–186.

45. Iyer, N. G., H. Ozdag and C. Caldas (2004). “p300/CBP and cancer.” Oncogene 23(24): 4225–4231.

46. Liu, X., L. Wang, K. Zhao, P. R. Thompson, Y. Hwang, R. Marmorstein and P. A. Cole (2008). “The structural basis of protein acetylation by the p300/CBP transcriptional coactivator.” Nature 451(7180): 846–850.

47. Qian, M., H. Zhang, S. K. Kham, S. Liu, C. Jiang, X. Zhao, Y. Lu, C. Goodings, T. N. Lin, R. Zhang, T. Moriyama, Z. Yin, Z. Li, T. C. Quah, H. Ariffin, A. M. Tan, S. Shen, D. Bhojwani, S. Hu, S. Chen, H. Zheng, C. H. Pui, A. E. Yeoh and J. J. Yang (2017). “Whole-transcriptome sequencing identifies a distinct subtype of acute lymphoblastic leukemia with predominant genomic abnormalities of EP300 and CREBBP.” Genome Res 27(2): 185–195.

48. Vo, N. and R. H. Goodman (2001). “CREB-binding protein and p300 in transcriptional regulation.” J Biol Chem 276(17): 13505–13508.

49. Bernt, Kathrin M., N. Zhu, Amit U. Sinha, S. Vempati, J. Faber, Andrei V. Krivtsov, Z. Feng, N. Punt, A. Daigle, L. Bullinger, Roy M. Pollock, Victoria M. Richon, Andrew L. Kung and Scott A. Armstrong (2011). “MLL-Rearranged Leukemia Is Dependent on Aberrant H3K79 Methylation by DOT1L.” Cancer Cell 20(1): 66–78.

50. Zhang, Y. and T. G. Kutateladze (2019). “Methylation of Histone H3K79 by Dot1L Requires Multiple Contacts with the Ubiquitinated Nucleosome.” Mol Cell 74(5): 862–863.

51. McGinty, R. K., J. Kim, C. Chatterjee, R. G. Roeder and T. W. Muir (2008). “Chemically ubiquitylated histone H2B stimulates hDot1L-mediated intranucleosomal methylation.” Nature 453: 812.

52. Anderson, C. J., M. R. Baird, A. Hsu, E. H. Barbour, Y. Koyama, M. J. Borgnia and R. K. McGinty (2019). “Structural Basis for Recognition of Ubiquitylated Nucleosome by Dot1L Methyltransferase.” Cell Rep 26(7): 1681–1690 e1685.

53. Jang, S., C. Kang, H. S. Yang, T. Jung, H. Hebert, K. Y. Chung, S. J. Kim, S. Hohng and J. J. Song (2019). “Structural basis of recognition and destabilization of the histone H2B ubiquitinated nucleosome by the DOT1L histone H3 Lys79 methyltransferase.” Genes Dev 33(11-12): 620–625

54. Valencia-Sanchez, M. I., P. De Ioannes, M. Wang, N. Vasilyev, R. Chen, E. Nudler, J. P. Armache and K. J. Armache (2019). “Structural Basis of Dot1L Stimulation by Histone H2B Lysine 120 Ubiquitination.” Mol Cell 74(5): 1010–1019 e1016.

55. Worden, E. J., N. A. Hoffmann, C. W. Hicks and C. Wolberger (2019). “Mechanism of Cross-talk between H2B Ubiquitination and H3 Methylation by Dot1L.” Cell 176(6): 1490–1501 e1412.

56. Yao, T., W. Jing, Z. Hu, M. Tan, M. Cao, Q. Wang, Y. Li, G. Yuan, M. Lei and J. Huang (2019). “Structural basis of the crosstalk between histone H2B monoubiquitination and H3 lysine 79 methylation on nucleosome.” Cell Res 29(4): 330–333.

57. Luo H, Zhu G, Xu J, Lai Q, Yan B, Guo Y, Fung TK, Zeisig BB, Cui Y, Zha J, Cogle C, Wang F, Xu B, Yang FC, Li W, So CWE, Qiu Y, Xu M, Huang S. HOTTIP lncRNA Promotes Hematopoietic Stem Cell Self-Renewal Leading to AML-like Disease in Mice. Cancer Cell. 2019 Dec 9;36(6):645-659.e8.

58. Yoshimoto, R., A. Mayeda, M. Yoshida and S. Nakagawa (2016). “MALAT1 long non-coding RNA in cancer.” Biochimica et Biophysica Acta (BBA) - Gene Regulatory Mechanisms 1859(1): 192–199.

59. Jianghua Liu, W.-X., Peng, Yin-Yuan Mo, Dianzhong Luo (2017). “MALAT1-mediated tumorigenesis.” Frontiers in Bioscience.

60. Sun, W., Y. Yang, C. Xu and J. Guo (2017). “Regulatory mechanisms of long noncoding RNAs on gene expression in cancers.” Cancer Genetics 216: 105–110.

61. Wei, Y. and B. Niu (2015). “Role of MALAT1 as a Prognostic Factor for Survival in Various Cancers: A Systematic Review of the Literature with Meta-Analysis.” Disease markers 2015: 164635–164635.

62. Zhao, M., S. Wang, Q. Li, Q. Ji, P. Guo and X. Liu (2018). “MALAT1: A long non-coding RNA highly associated with human cancers.” Oncology letters 16(1): 19–26.

63. Li, Z. X., Q. N. Zhu, H. B. Zhang, Y. Hu, G. Wang and Y. S. Zhu (2018). “MALAT1: a potential biomarker in cancer.” Cancer Manag Res 10(1179-1322 (Print)): 6757–6768.

