## Supplementary Figure for "Prognostic significance of protein-coding and long non-coding RNA expression profile in T-cell acute lymphoblastic leukemia"

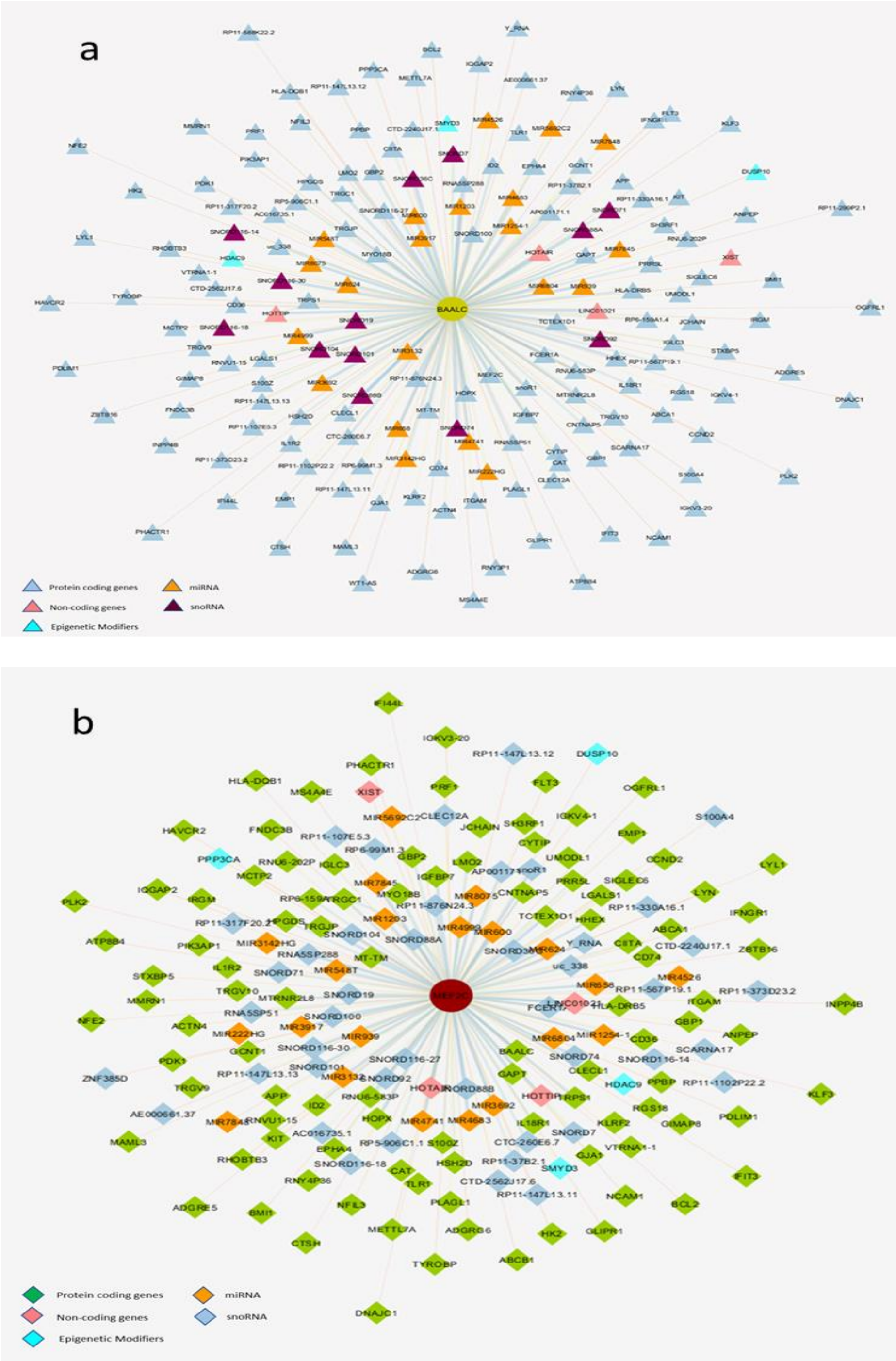


**Supplementary Figure 1:** Enriched gene ontology functions of deferentially expressed genes, DAVID results the number of involved genes from differentially expressed genes in T-ALL for biological process **(a)**, cellular components **(b)** and KEGG pathways**(c).** (p<0.005). Complete list of genes involved in the biological process and cellular component of are shown in Supplementary

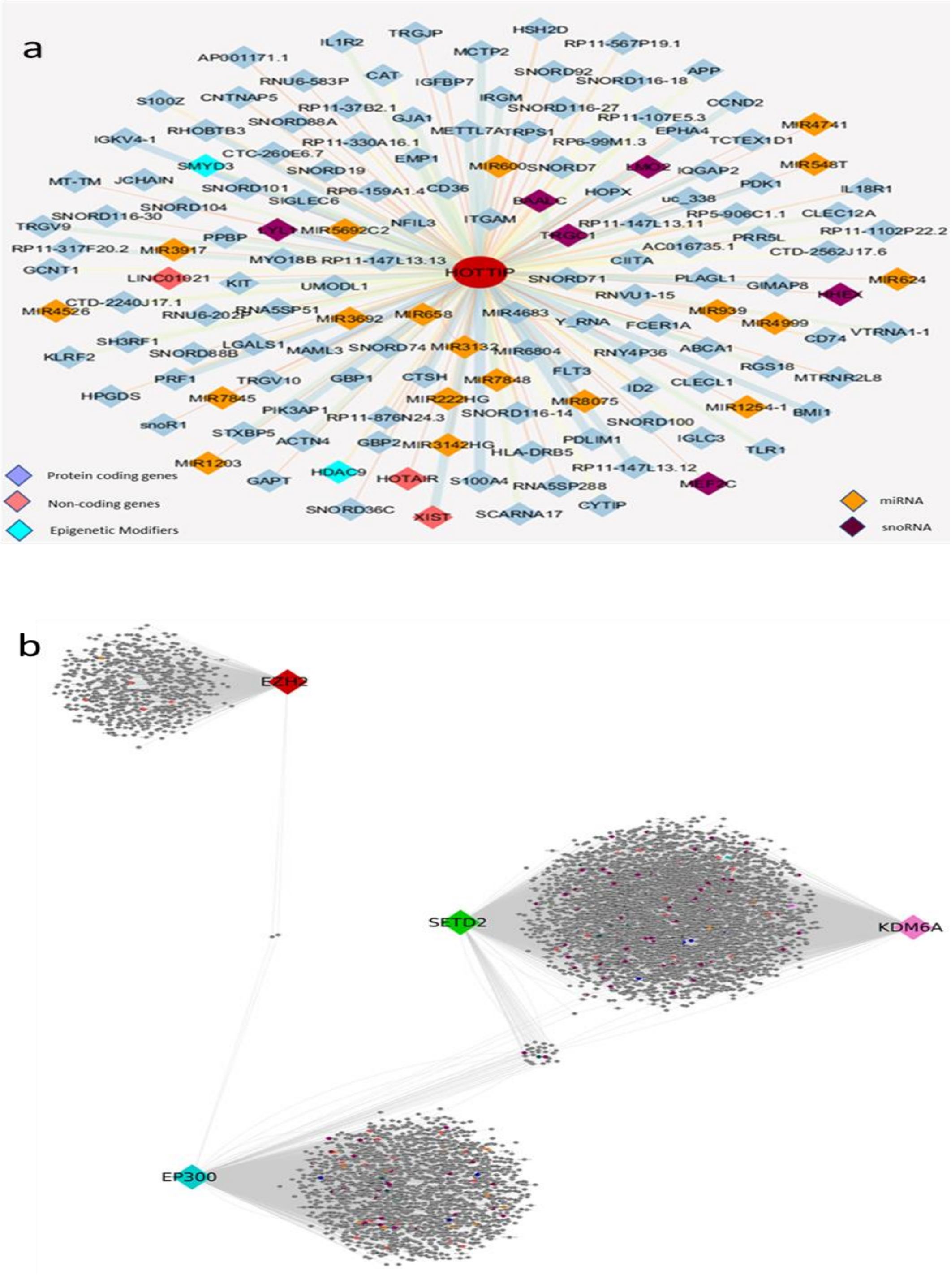


**Supplementary Figure 2: (a)** Co-expression network profile of *BAALC* correlated genes Cytoscape network visualizing the immature group. **(b)** Co-expression network profile of *MEF2C* correlated genes Cytoscape network visualizing the immature group.
