## Supplementary Method for "Prognostic significance of protein-coding and long non-coding RNA expression profile in T-cell acute lymphoblastic leukemia"

### Discovery Cohort:

A total of 35 T-ALL patients based on morphology, cytochemistry and immunophenotypic features were studied for RNA-Seq. The age of the patients ranged from 1 to year 55 with a median age of years. There were 26 males (19 pediatric, 7 adults) and 9 females (7 pediatric, 2 adults). The mean hemoglobin (Hb), total leukocyte count (TLC) and platelet count (Plt) was 8.5 gm/dl, 166,300/µL and 54,500/µL, respectively. The immunophenotypic subtype of these patients is shown in Table 1 . Transcriptome data from a pool of total RNAs from 5 normal human thymuses were used as control. (this was kindly provided by Prof. Jan Cools, Leuven, Belgium)

**Table 1: Distribution of T-ALL patients in discovery cohort based on immunophenotype**

| **Subtype of T-ALL** | | **Number of patients** |
|---|---|---|
| Immature T-ALL | ETP-ALL | 5 |
| | Non ETP-ALL | 8 |
| Cortical T-ALL | | 17 |
| Mature T-ALL | | 5 |

### Morphological diagnosis

In this study, the morphological evaluation was performed by hematopathologist, in accordance with (Vardiman, Thiele et al. 2009, Taylor, Xiao et al. 2017). For each case, routine well-prepared

Jenner-Giemsa-stained smears were evaluated. Cytochemistry for myeloperoxidase (MPO) was done in each case.

**Flow cytometric analysis**

All monoclonal antibodies were obtained from Beckman Coulter [BC], Hialeah, FL. The antibodies were conjugated to fluorescein isothiocyanate (FITC), phycoerythrin (PE), PE-Texas Red (ECD), PE-cyanin 5 and PE-cyanin 7 (PE-Cy7). The antibodies :*CD3, CD45, CD2, CD7, CD4, CD5, CD8, CD1a, CD13, CD33, CD117, HLA-DR, CD34, CD65* and *CD11b* were used in flow cytometry. All procedure, sample acquisition and analysis protocol were used as described in publication from our group.(Chopra, Bakhshi et al. 2014).

The **s**ubclassification of patients was done based on EGIL classification.(Bene, Castoldi et al. 1995). ETP-ALL immunophenotype was characterized by an absence (<5% positive lymphoblasts) of *CD1a*, *CD8* expression , weak *CD5* expression with less than 75% positive lymphoblast, and expression of one or more of the myeloid or stem cell markers (*CD117, CD34, HLA-DR, CD13, CD33, CD11B or CD65*) on at least 25% of lymphoblast (Coustan-Smith, Mullighan et al. 2009).

***RNA* sequencing**

PBMC isolation performed by density gradient method using Histopaque (Sigma-Aldrich, USA) and 1 ml TRIzol (Thermo Fisher Scientific, USA) for per one million cells was added in each vial immediately and mixed by syringing. TRIzol mixed samples were stored at -80°C for RNA isolation. RNA was isolated by TRIzol (Guanidinium thiocyanate-phenol-chloroform extraction) method (Rio Dc Fau - Ares, Ares M Jr Fau - Hannon et al.). RNA was dissolved in DEPC-treated waterh a pipette tip. RNA quantity was checked by nanodrop and Qubit RNA broad range assay kit on dye-based qubit spectrophotometer (Thermo Fisher Scientific, U.S.). Agilent 2100 Bioanalyzer (Agilent Technologies, USA) with RNA chips was used to check the RNA Integrity Score (RIN). We selected samples with ≥ 7 RIN score for RNA sequencing library preparation.

### RNA seq library preparation

Sample preparation for sequencing was carried out using strand specific Truseq RNA sample preparation kit (Illumina, San Diego, California, U.S.) as per supplier's instructions and 8.0 picomol of the pooled library was sequenced on the Illumina HiSeq2000 system (Figure 1 ).

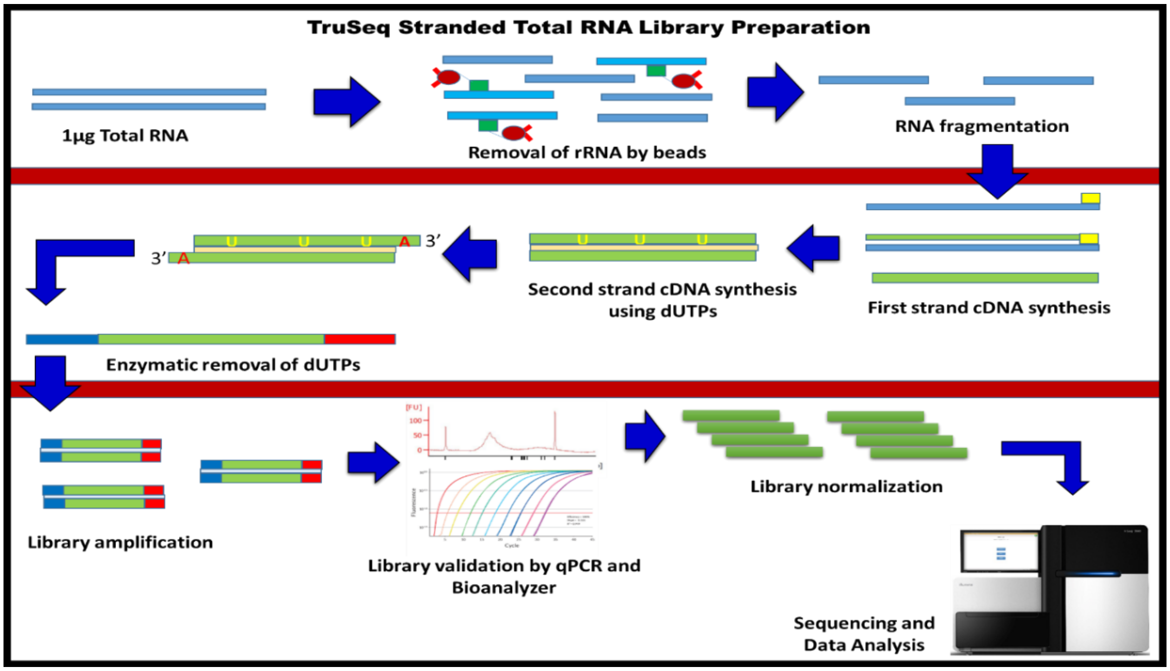

**Methods: Schematic diagram of library preparation by using Illumina TruSeq Stranded Total RNA library preparation kit**

### Data collection and Quality analysis

Total of 480 GB raw data was collected in the form of ". fastq" file which contained 50 million average reads per file. The quality of reads was checked using FastQC v0.11.8 (Andrews 2010) and trimming were implemented using Trimmomatic PE (Bolger, Lohse et al. 2014). We used a quality filter of Phred quality cut-off 30 and ahead crop of 15. The overall percentage of reads surviving after Trimmomatic was 70.83% to 86.36%.(Figure 2&3)

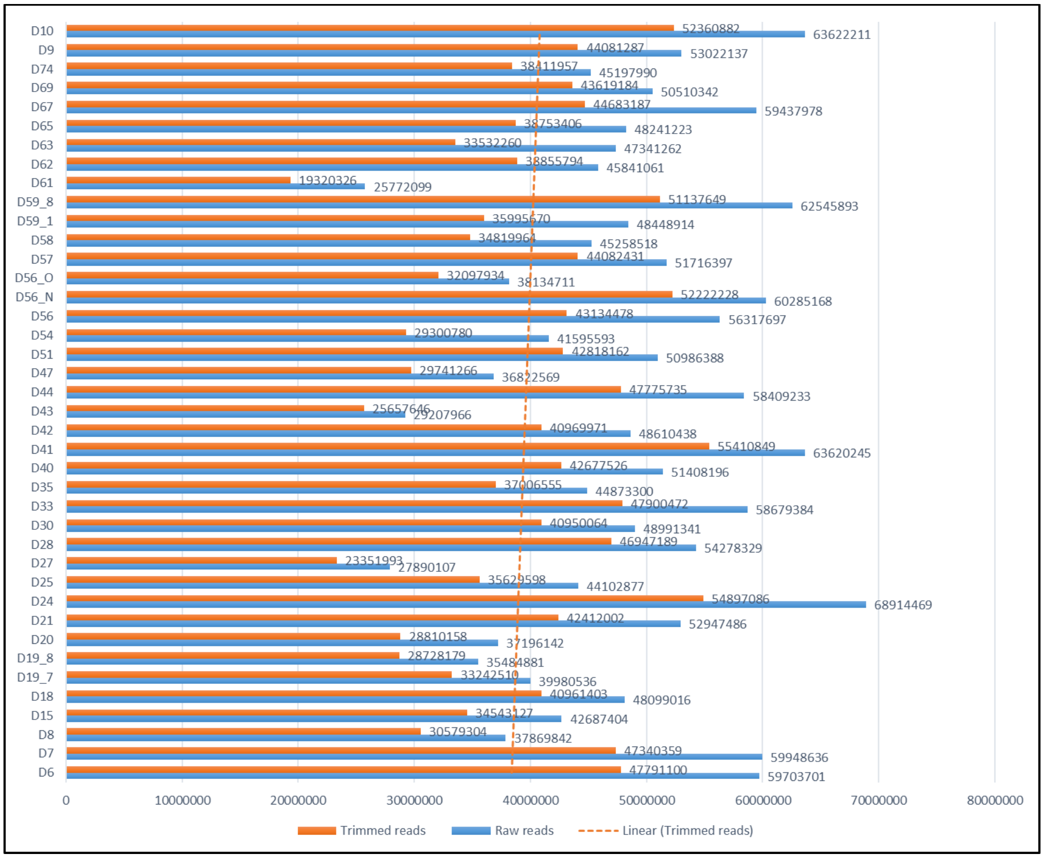

**Figure 2 : Graphical representation of raw and trimmed reads**

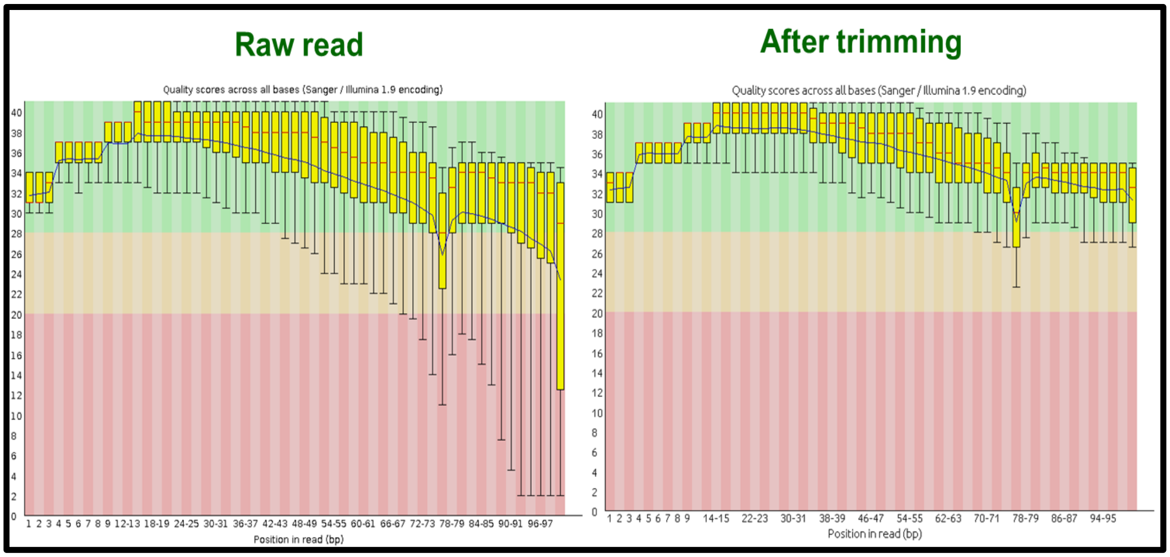

**Figure 3: Quality check of sample D6 raw reads and after running Trimmomatic software by FastQC.**

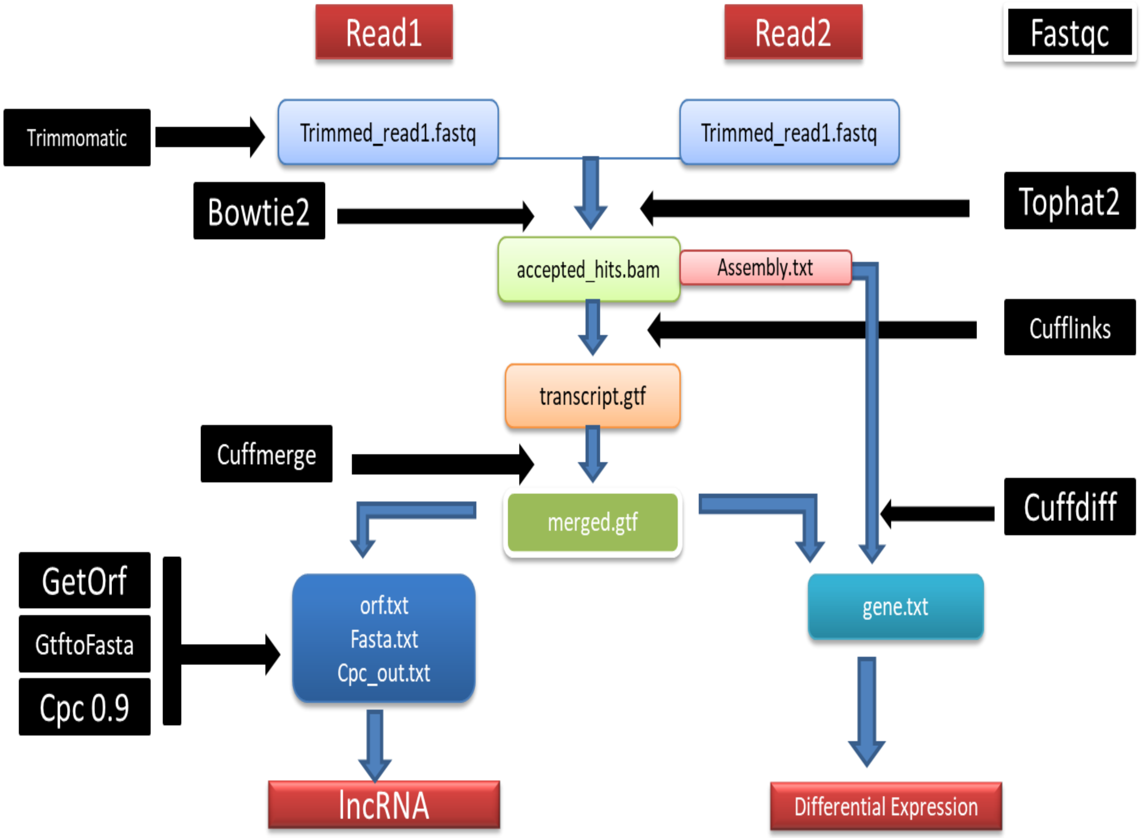

**Figure 4: Data analysis pipeline for differentially expressed coding genes and long non-coding RNA**

### Mapping and alignment of reads

Brief analysis pipeline was described in Figure 4. Human reference genome, hg38, was used for mapping and alignment, downloaded from UCSC genome browser gateway. Reads were aligned to the indexed hg38 genome using splice junction mapper: Tophat2.1.1 (Trapnell, Pachter et al. 2009, Kim, Pertea et al. 2013). Total Avg 82.275 % reads were aligned to the human genome hg38.

### Transcriptome assembly and merging:

Cufflinks 2.2.1 (Trapnell, Williams et al. 2010) was used to assemble the transcripts, estimate transcripts abundance and calculate differential gene expression and regulation. These assemblies .gtf files are then merged together into a single file by using the Cuffmerge utility. Finally, merged files were stored in merged.gtf format.

### Differential gene expression and functional analysis by the supervised approach

Differential expression of the assembled transcripts across 3 types of T-ALL (immature, cortical and mature T-ALL based on immunophenotypic features) were analyzed using module Cuffdiff from the Cufflinks package (Trapnell, Williams et al. 2010). Samples of cortical subgroup were also analyzed in sCD3 positive and negative conditions by Cuffdiff. The Fragment per Kilobase Exon per Million Reads (FPKM) measure was used to quantify the expression of the transcript. For better understanding, these FPKM values were converted into fold changes. We filtered in all genes which were ≥ 2-fold changes as a significantly important gene for a given condition.

Gene functional analysis was done on DAVID 6.8 database (Huang da, Sherman et al. 2009, Huang da, Sherman et al. 2009). Gene list of each subgroup was analyzed and result for "biological process, cellular component and KEGG pathways were filtered in, based on p-value $\leq$0.005.

**Differential gene expression analysis by the unsupervised approach**

The output of cufflink was used in cuffnorm to produce several output files that contain normalized fragment counts and expression levels at the level of transcripts, primary transcripts, and genes. In this approach, 35 patients and transcriptome from normal thymus was used. We found 58,224 genes that were expressed in all 35 patient samples including normal thymus. In cuffnorm output, we removed all transcripts which were < 2 FPKM scores and we found 31,694 genes that were $\geq$2 fpkm. All samples were used as an input feature to perform principal component analysis (PCA) on BioVinci Version: 1.1.5, r20181005 (Bioturing, USA). We found maximum principle component equal to the samples and only 6 were statistically significant. To apply an unsupervised approach all 6 principal components were analyzed, and a graph was plotted. In this graph, all sample were clustered into 3 major clusters. After this, all samples were compared with immunophenotypic phenotypes of samples.

**Differential expression analysis of non-coding RNA**

**Known lncRNA**

The long noncoding RNAs were identified and characterized by "Biomart" (https://www.ensembl.org/biomart). Later their differential gene expression analysis was performed. All transcript which had more then 1FPKM were used to calculate fold changes. Transcripts those were more than 2-FC and less than -2 FC was selected as a putative transcript in a given condition. The results were plotted on heatmap using multiple experiment viewer (MeV, http://mev.tm4.org). Further, their gene function co-expression network analysis was performed.

### Novel lncRNA

### Annotation of novel long non-coding RNA

For identification of novel lncRNAs, the resulting output of cuffmerge was used to analyze lncRNA in the data. The transcripts with a nucleotide length greater than 200 were filtered. These transcripts were further classified as coding and non-coding by evaluating the length of their open reading frames (ORF) and coding potentials using two independent approaches, ORF finder and CPC calculator (Kong, Zhang et al. 2007). The ORFs of all the transcripts were predicted using getORF utility of EMBOSS toolkit. Transcripts having an ORF of length greater than or equal to 30 amino acids (aa) were removed in order to remove all protein-coding as well as micro peptide coding transcripts, thus giving more stringent criteria to predict only noncoding transcripts as lncRNAs. The coding potential of the filtered transcripts was further calculated using the Coding Potential Calculator (CPC). A CPC scores less than zero indicates a low coding potential of a transcript while positive CPC score indicates a high coding potential of a transcript. The transcripts with a score less than 0 were retained and the final set represented the long non-coding RNAs of zebrafish while the one with orf more than30 aa and CPC score of more than 1 were considered as protein-coding transcripts. The pipeline used in the analysis of novel lncRNA has been summarized in Figure 4.

Differential expression analysis of lncRNAs: Differential expression of the assembled transcripts across various types of T-ALL (immature, cortical and mature T-ALL) was analyzed using Cuffdiff. The FPKM measure was used to quantify the expression of lncRNAs. lncRNAs which have at least >5 FPKM in one type of T-ALL and at least a 2-fold difference between other conditions were prioritized for further validation.

### Differential gene expression of small non-coding RNA (miRNA)

Like, lncRNAs, miRNAs were identified from "Biomart" (https://www.ensembl.org/biomart). To get the more information differential gene expression analysis was performed. All transcripts which were ≥ to 1 fpkm were used to calculate fold changes. Transcripts with ≥ 2-FC were selected as putative miRNA transcript in given subgroup and heatmap was plotted using MeV. Due to insufficient sample volume, real-time validation could not be performed on RNA-Seq samples for miRNA.

### Gene co-expression network analysis

In order to predict the functions of the genes, histone modifiers, lncRNA, miRNA in all three subgroups a correlation network analysis was performed. A correlation matrix was generated using Pearson correlation as a statistical measure using R scripts based on the FPKM scores of the genes and package Hmisc where rcorr function was used. A threshold of "r" value 0.9 and "p-value" less than 0.05 was used to construct the co-expression network. Cytoscape (Shannon, Markiel et al. 2003) was used to visualize the network where rscore was used as attribute measure. All DF genes, lncRNA, miRNA and histone modifiers were used as a source node and other genes as target source. The 3D-way layout was used for visualization of this network.

### Data visualization

All differentially expressed gene in all the subgroups were visualized by heatmap based on the log scale and hierarchical clustering based on complete linkage as input settings. For gene network analysis, Cytoscape tool (Shannon, Markiel et al. 2003) was used that allows to build a correlation network in between the gene and other entity based on r and p-value. In network analysis, all gene nodes were plotted using a 3D layout.

### Validation of RNA seq results on the discovery cohort

Validation of data was done by reverse transcriptase PCR followed by real-time PCR using specific primers and probes listed in Table 2 & 3 Primer3 (Soulier, Clappier et al. 2005).

cDNA synthesis was done using Vilo reverse transcriptase kit (Thermo Fisher Scientific, USA) with 100ng of RNA. Further cDNA was diluted in 1:10 ration in nuclease-free water.

**Table 2: Primer sequence used for validation by real-time PCR.**

| | | **Forward primer (5'-3')** | **Reverse (5'-3')** |
|---|---|---|---|
| **Immature** | *MEF2C* | GCGCTGATCATCTTCAAC | CTTTGCCTGCTGCTGATCATT |
| | *BALLC* | GCCCTCTGACCCAGAAACAG | CTTTTGCAGGCATTCTCTTAGCA |
| | *LYL1* | TGGCCCTGCACTACCACC | AGATGCTAAAAGGTCCTGCTGG |
| | *LMO2* | CTGCCTGAGCTGCGACCT | GTTTGTAGTAGAGGCGCCGC |
| | *HHEX* | TCTGCATAAAAGGAAAGG | CGTCTCGAATTTCTTCTC |
| | *TRGC1* | TACCTCCTCCTGCTCCTCAA | GAGAACTGAAATGGCCCAAA |
| House keeping | *ABL1* | TGGAGATAACACTCTAAGCATAACTAAAGGT | GATGTAGTTGCTTGGGACCCA |
| | *GAPDH* | GAAGGTGAAGGTCGGAGTCAAC | CAGAGTTAAAAGCAGCCCTGGT |
| | *β-actin* | TCACCCACACTGTGCCCATCTACGA | CAGCGGAACCGCTCATTGCCAATGG |
| | *HOX11 (TLX1)* | TGGAGAGTAACCGCAGATACACA | CGTGCGCGGCTTCTTC |
| | *TRGV2* | CCTCAGTCACTTCCCTCTGC | GACTGGCAGGAGACAGGAAA |

| | | | |
|---|---|---|---|
| Cortical | *TLX3* | ACGGTCTCCAGCCTTGGC | GCGCCGGGTCACGGT |
| | *FAT1* | TGCTACAGACGCAGACATCC | AACAGCTTGCTCCTCACGAT |
| | *MEG11* | GGAGCTGAATCCCTACACCA | GTCTCGGCCCTTCTCTTTCT |
| | *LMO1* | CAACGTGTATCACCTCGACTGC | GAAGAATTTGTCTCCCACACAAAA |
| | *FGR* | GCATCCAAGGTGTGGAGTTT | GCAACGAAGGGGACATTTTA |
| | *TTK* | CAGCAGCAACAGCATCAAAT | TGCTTGAACCTCCACTTCCT |
| Mature | *ST20* | CTCTGTCTCCCAGGTTCAGG | TGCTTCAGGACAGAAGAGCA |
| | *EML4* | CCATGCAACGAGATAAGCAA | CACATGCAGCTGAAGGAAAA |
| | *CDCA2* | GCCCTGCACTGTATCGAAAT | CGCTGAGACCTTCCTTTCTG |
| | *TAL1* | TTCACCACCAACAATCGAGTG | TGCCGCCATCGCTCC |
| | *CREG2* | TCTCTGCCTCCTGATCCTGT | GGCAAAGTCCTTTGAAGCAG |
| lncRNA | | | |
| | *XIST* | AAGCTAAGGGCGTGTTCAGA | TGACTTCCTCTGCCTGACCT |
| | *LINC01221* | TTCCAGGCAAATCCTTTTTG | ACCAGAGAAGCCTGCCAGTA |
| | *TRBV11-2* | CTCCTGGGAGCAGAACTCAC | TAAAGGGTAGCATGGCCAGA |
| | *MALAT1* | TGTGTGCCAATGTTTCGTTT | AGGAGAAAGTGCCATGGTTG |
| | *CYTOR* | CGGAATGCAGCTGAAAGATT | ACCAGCCCATGACCAAAATA |
| | *HOTAIR* | AAGGCCCCAAAGAGTCTGAT | AGCACTTCTCTCGCCAATGT |
| | *HOTTIP* | AAGGGTCTCAGCTCCACAGA | CTGCCGTCTTTTCTGAGTCC |
| Epigenetic | *RAG1* | AGCCTGCTGAGCAAGGTACC | GAACTGAGTCCCAAGGTGGG |
| | *EZH2* | TTCATGCAACACCCAACACT | GAGAGCAGCAGCAAACTCCT |
| | *KDM6A* | CCTCATAACCGCACAAACCT | CTGCCGAATGTGAACTCTGA |
| | *EP300* | CAAACGCCGAGTCTTCTTTC | GTTGAGCTGCTGTTGGCATA |
| | *SETD2* | TCACAAGGCAGACTCAGTGG | CTGCTGTCTTGGGCTTTTTC |

### Table 3: Probe sequence for real time validation

| ***GENE*** | **5'-PROBE-3'** |
|---|---|
| *MEF2C* | FAM- CAAGCTGTTCCAGTATGCCA-BHQ1 |
| *LYL1* | FAM-TCACCCCTTCCTCAACAGTGTCTACATTGG-BHQ1 |
| *HHEX* | FAM-AGATTCTCCAACGACCAGACCATC-BHQ1 |
| *TRGC1* | TET-AGAAGAACGGCTTTCTGCTG-BHQ1 |
| *β-actin* | QUASAR705-ATGCCCTCCCCCATGCCATCCTGCGT-BHQ1 |
| *HOX11(TLX1)* | TET-AGGACAGGTTCACAGGTCACCCCTATCAGAA-BHQ1 |
| *TRGV2* | CY5-TGCTCCTCTTCATCTGGTCC-BHQ1 |
| *FAT1* | TxRED-ACGTTATTGGGTTCAGGTGC-BHQ1 |
| *MEG11* | TET-CAGGCATCATGCTCCTGTTA-BHQ1 |
| *FGR* | CY5-CTGCTCATCTGCACGTGAGT-BHQ1 |
| *TTK* | TET-CAGCAAATGAATGCATTTCG-BHQ1 |
| *ST20* | TxRED-GCCTAAATCTGGCTGGATGA-BHQ1 |
| *EML4* | TxRED-GTCCTAACACCCTGGCTTCA-BHQ1 |
| *CDCA2* | FAM-CTGGAATTCTCAGAGGCAGG-BHQ1 |

| | |
|---|---|
| *TAL1* | TET-ATTACTGATGGTCCCCACACCAAAGTTGTGC-BHQ1 |
| *CREG2* | CY5-TAAGGTCCCCTGGCTAGGTT-BHQ1 |
| *XIST* | TET-CCATTTTAGACTTGCCGCAT-BHQ1 |
| *LINC01221* | TXRED-CAGTCTGAACAGCCCAGTGA-BHQ1 |
| *TRBV11-2* | CY5-GAGAAAAGGCAGAGTGTGGC-BHQ1 |
| *MALAT1* | QUASAR705-CAGCAGGCAGCTGTTAACAG-BHQ1 |
| *CYTOR* | TXRED-CTGTGGACTCTGAGGCCTCT-BHQ1 |
| *HOTAIR* | CY5-AAAGGCTGAAATGGAGGACC-BHQ1 |
| *HOTTIP* | CY5-GAGGAGCCACAAGCAGGTAC-BHQ1 |
| *EZH2* | CY5-TTACCAGCATTTGGAGGGAG-BHQ1 |
| *KDM6A* | TXRED-AGCCGTGGAAAAACCAACTA-BHQ1 |
| *EP300* | TET-TCAATGTTGCATTCAGCCAT-BHQ1 |
| *SETD2* | QUASAR705-TGAGATGGACCTGGGAACTC-BHQ1 |

### Validation cohort:

#### Patients characteristics

A total of 99 T-ALL patients including 70 children and 29 adults were studied for validation of RNA-Seq results. The age of the patients ranged from 3 to 65 years with a median age of 12 years. There were 86 males (61 pediatric, 25 adults) and 13 females (9 pediatric, 4 adults). The mean hemoglobin (Hb), total leukocyte count (TLC) and platelet count (Plt) was 9.5 gm/dl, 104,922/µL and 87,895/µL, respectively. The immunophenotypic subset of these patients are shown in Table 1 of the 99 T-ALL patients in the validation cohort, RNA in enough quality and quantity was available for the determination of MEF2C in 99 and BAALC, HHEX, LYL1, TAL1, FAT1, XIST genes in 87 cases.

**Table 3: Distribution of T-ALL patients in validation of based on immunophenotype into immature, cortical and mature subtypes.**

| **Subtype of T-ALL** | **Number of patients** |
|---|---|

| | | |
|---|---|---|
| Immature T-ALL | ETP-ALL | 13 |
| | Non ETP-ALL | 32 |
| Cortical T-ALL | | 44 |
| Mature T-ALL | | 10 |

The results of RNAseq data were validated in the validation cohort using realtime PCR to assess its clinical and prognostic significance.

### Treatment

In the discovery cohort, 14 patients were treated with ICICLE protocol, 3 with Berlin-Frankfurt-Munster-90 (BFM-90) protocol, 1 with INCTR, 1 with Holzer's protocol and 3 with hyper CVAD. 13 patients did not take treatment.

In the validation cohort, seventy-two patients were treated with ICICLE protocol, 15 with Berlin-Frankfurt-Munster-90 (BFM-90) protocol and 3 with hyper CVAD. Nine patients did not take any treatment. Two patients died during induction chemotherapy. Complete remission was defined as bone marrow blasts <5% with a recovery of blood counts at the end of 4 weeks of induction chemotherapy. Any failure to do so (including the persistence of leukemic blasts in an extramedullary site), or death during induction therapy due to any cause, was considered as induction failure Patients who failed with one protocol were re-inducted with another.

### Statistical analysis

Fisher's exact test for categorical data was used to compare baseline clinical variables across groups in the validation cohort. A P value $\leq$0.05 (two-sided) was considered significant. *BAALC*, *LYL1, FAT, TAL1* and HHEX were dichotomized at its median with patients classified as low gene expression if they had expression values within the lower 50% and as high gene expression if they had gene expression values within the upper 50%. For *MEF2C* gene expression, the patients were dichotomized into low *MEF2C* expression and high *MEF2C* expression based on a cutoff of 0.1- *MEF2C* high, if the expression is $\geq$0.1 and *MEF2C* low if the expression is <0.1. This cut-off was chosen based on our unpublished data from a larger number of patients, which showed that the patients can be divided into groups with clinically distinct event-free survival and overall survival. For *XIST* gene expression, we took the cut-off of 1. Since the *XIST* gene is present on X-chromosome, we did analysis for all the patients and only male patients separately, to see the association of various parameters and *XIST* gene expression.

Response to prednisolone was assessed by the examination of peripheral blood at day 8 of initiation of prednisolone therapy. The patients were divided into prednisolone sensitive and resistant based on the presence of <1000/µL and $\geq$1000/µL blasts, respectively.

Achievement of complete remission (CR) was assessed after completion of induction chemotherapy and presence of absolute neutrophil count >1000/µL, platelet count >100,000/µL and hemoglobin >10gm/dL, no peripheral blood blasts, less than 5% BM blasts and absence of extramedullary leukemia. Relapse was defined as the reappearance of blasts in the peripheral blood, more than 5% BM blasts, or development of extramedullary leukemia.

The median follow-up was 16 months. After excluding 13 patients in validation cohort who did not receive treatment, 90 patients were included for the analysis of event-free survival and overall survival.

Event-free survival (EFS) was defined as the time from diagnosis to the date of the last follow-up in complete remission or the first event (i.e., induction failure, relapse, secondary neoplasm, or death from any cause). Failure to achieve remission due to non-response was considered an event at time zero. Survival was defined as the time from diagnosis to death or the last follow-up. Patients lost to follow-up were censored at the last contact. The last follow up was carried out on April 2019. The Kaplan-Meier method was used to estimate survival rates, with the differences compared using a two-sided log-rank test. Cox proportional hazard models were constructed for EFS and OS and used for univariate and multivariate analyses. Covariates included in the full model of OS and EFS were Sex, WBC (<100X $10^9$/L, >100X$10^9$/L), and age (<10 years vs. >10 years), gene expression, immunophenotype, CNS involvement, response to prednisolone treatment, bone marrow remission status and presence of minimal residual disease after the end of induction chemotherapy. All analyses were performed using the SPSS statistical software package, version 20.0/STATA software, version 11.
